# Development and design of the BELpREG registration system for the collection of real-world data on medication use in pregnancy and mother-infant outcomes

**DOI:** 10.1101/2023.03.01.23286625

**Authors:** Laure Sillis, Veerle Foulon, Karel Allegaert, Annick Bogaerts, Maarten De Vos, Titia Hompes, Anne Smits, Kristel Van Calsteren, Jan Y Verbakel, Michael Ceulemans

## Abstract

Although medication use during pregnancy is common, most available products lack sufficient safety information. As prospective data collection and perinatal pharmacoepidemiologic research on medication safety in pregnancy did not exist in Belgium yet, the BELpREG data registration system was developed. BELpREG enables comprehensive ‘real-world’ data collection on perinatal medication use and mother-infant outcomes via online questionnaires that are completed by pregnant women every four weeks during pregnancy and in the first eight weeks after childbirth. This paper describes the development and current design of the BELpREG system, including the list of BELpREG variables. To compile this list of variables, relevant documents were explored, followed by consultation of an interdisciplinary expert panel. The included variables were structured in seven categories: 1) Sociodemographic characteristics; 2) Information on the current pregnancy and health status; 3) Maternal-obstetric history; 4) Use of medicines, folic acid / pregnancy vitamins and other health products; 5) Substance use; 6) Pregnancy outcomes; and 7) Neonatal outcomes. An electronic informed consent and linkage to medication databases, with images of drug packages and underlying structured data fields, are built into the system. Data collection has officially started in November 2022. Based on its rigorous design, BELpREG holds the potential to be a successful and sustainable research tool, enabling perinatal pharmacoepidemiologic research in Belgium and beyond.

## 1 Introduction

The use of medicines and health products in pregnancy occurs frequently (Lupattelli et al., 2014; Gerbier et al., 2021; Ceulemans et al., 2022a; Gerbier et al., 2022). In Belgium, almost 90% of the women use at least one medicine in pregnancy, excluding pregnancy vitamins (Larcin et al., 2021). Safe use of medication in pregnancy is vital as the exposure to certain medicines may result in adverse pregnancy, neonatal and/or infant outcomes. To date, there is a high need for reliable safety information on perinatal medication use, also in Belgium (Ceulemans et al., 2020; Ceulemans et al., 2022b). The Internet, including social media, is a commonly used source to search for safety information (Sinclair et al., 2018), however, discrepancies have been found between online sources (Nörby et al., 2021). Furthermore, social media posts often provide inaccurate information, affecting women’s perception and decisions about medication use (van Gelder et al., 2019).

Despite their frequent use, most available medicines lack sufficient safety information on their use during pregnancy (Adam et al., 2011). On average, 27 years are needed to assign a precise risk category to a medicine (Adam et al., 2011), and research to gain more information on the safe use of medicines in pregnancy is challenging. Pregnant women are commonly excluded from clinical trials assessing the efficacy and safety of new medicinal products (Shields and Lyerly, 2013; Scaffidi et al., 2017; Zhao et al., 2022). As soon as a product is on the market, marketing authorisation holders (MAHs) are involved in the continuous monitoring of medication safety in pregnancy via post-marketing surveillance, including spontaneous reporting and (mandatory) product-specific pregnancy registries. Still, a recent pan-European qualitative analysis performed by our team highlighted the difficulties with respect to data collection on this topic MAHs suffer from, including underreporting, the collection of incomplete information, and loss to follow-up (Sillis et al., 2022). A recent landscape analysis also confirmed that only few post-marketing studies result in label updates (Roque Pereira et al., 2022).

In addition to post-marketing surveillance, other data sources could be used to gain insight into medication safety during pregnancy, as for example: population-based cohorts nested in health utilisation databases (i.e., secondary use of health data) (Dandjinou et al., 2019; Huybrechts et al., 2021; Moseholm et al., 2022), pregnancy registries held by non-MAHs (with or without a specific focus on a pharmacotherapeutic class or therapeutic indication) (Hernandez-Diaz et al., 2012; Chambers et al., 2019; Vorstenbosch et al., 2019), and data collection by Teratology Information Services (Pauliat et al., 2021; Dao et al., 2022). Different data sources come with different strengths, limitations, and methodological considerations (Hernandez-Diaz and Oberg, 2015; Huybrechts et al., 2019; Panchaud et al., 2022). In 2019, the Innovative Medicines Initiative (IMI) project ConcePTION was launched to create a sustainable ecosystem to generate and disseminate information on medication safety in pregnancy and during breastfeeding. This goal also includes the alignment of different organisations and approaches aimed at improving in the field of medication safety for pregnant and breastfeeding women (ConcePTION, n.d.).

Unfortunately, in Belgium, pharmacoepidemiologic research on medication safety in pregnancy is lacking. The required data for such research are neither routinely collected, nor easily accessible, compared to other European countries with comprehensive data on birth cohorts (e.g., Norway, the MoBa cohort) (Magnus et al., 2006) or with linked health utilisation databases (e.g., France, EFEMERIS/POMME) (Lacroix et al., 2009; Benevent et al., 2019). This clearly highlights the need for comprehensive data registration in Belgium, and initiatives to expand this research field.

In 2021, our interdisciplinary consortium received KU Leuven funding to develop, validate and implement the BELpREG registration system in Belgium (“a **BEL**gian interdisciplinary initiative to enhance **p**regnancy related data **REG**istration and research on medication use”). The BELpREG registration system enables the collection of ‘real-world’, prospective, observational data on perinatal medication use and mother-infant outcomes, by using online questionnaires to be completed by pregnant women. BELpREG is a non-disease, indication, pharmacotherapeutic class or product-specific registry. In BELpREG, medication exposure and outcome data of all pregnant women (≥18 years and receiving health care in Belgium) are collected. The obtained data will be used for pharmacoepidemiologic research in Belgium and beyond. On the long-term, BELpREG aims to contribute to the provision of evidence on medication safety to expecting parents and HCPs. Figure 1 shows a schematic overview of the BELpREG approach.

**Figure 1.**
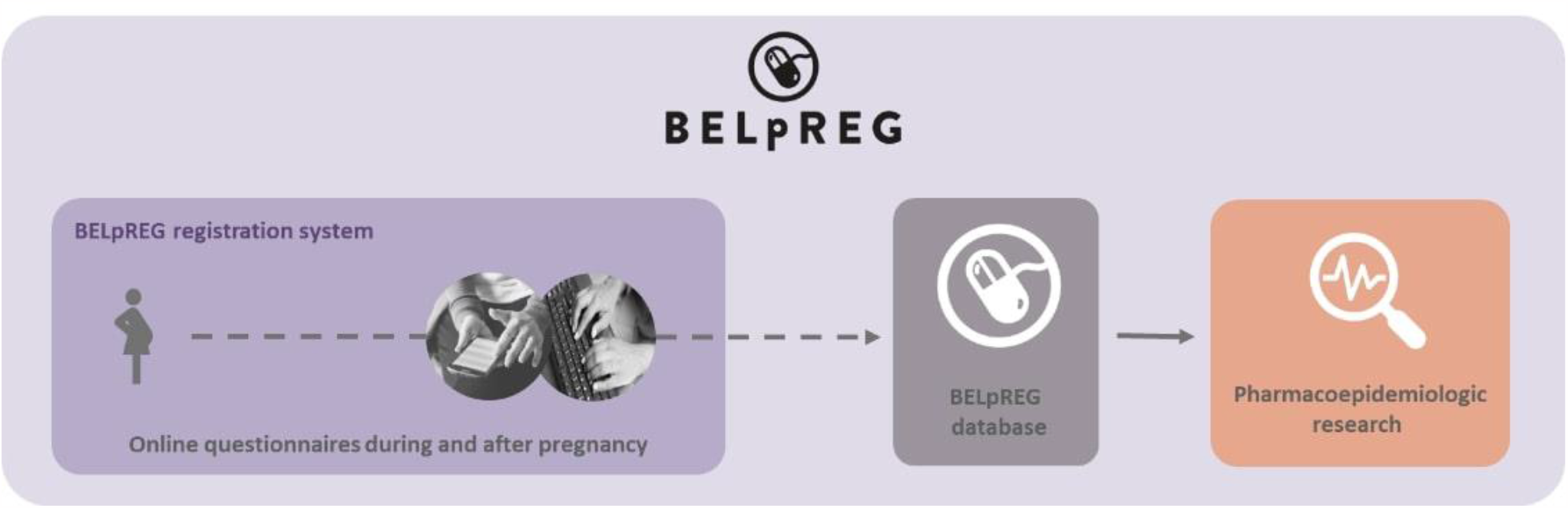
A schematic overview of the BELpREG approach.

In this paper, we describe the development and current design of the BELpREG registration system, including the list of BELpREG variables, and future possibilities. Hence, this overview will inform (inter)national researchers, MAHs, regulators and policy organizations involved in medication safety monitoring in pregnancy about the BELpREG initiative, germinating opportunities for international data pooling, analysis and collaborative projects.

## 2 Materials and Methods

### 2.1 Timing of the Development of BELpREG

The development of the BELpREG data registration system on perinatal medication use and mother-infant outcomes occurred between March 2021 and October 2022.

### 2.2 Development of the BELpREG Data Variables List

To compile the list of variables for data collection in BELpREG, relevant documents were explored, followed by a consultation of an interdisciplinary expert panel.

First, the list of Core Data Elements (CDE), which was recently compiled as part of the IMI ConcePTION project (version December 2021, available via https://www.imi-conception.eu/results/deliverables/) was consulted. The CDE had previously been defined based on in-depth discussions with experts in perinatal pharmacovigilance from in and outside the pharmaceutical industry, resulting in a set of essential variables required for pregnancy pharmacovigilance using prospective reports. The CDE aims to strengthen efficient collection of real-world data on medication safety in pregnancy (ConcePTION, n.d.). Second, the CDE were supplemented with variables defined in the European Medicines Agency (EMA) “Guideline on good pharmacovigilance practices (GVP) - Product- or Population-Specific Considerations III: Pregnant and breastfeeding women”. This document provides guidance to MAHs and competent authorities with respect to data collection on pregnancy exposures and outcomes by listing questionnaire elements (see Appendix 1 of the GVP guideline, version December 2019, available via https://www.ema.europa.eu/en/documents/scientific-guideline/draft-guideline-good-pharmacovigilance-practices-product-population-specific-considerations-iii_en.pdf). Finally, additional input regarding the variables and response options was sought in questionnaires of existing registration systems (i.e., Moeders van Morgen, The Netherlands; MotherToBaby, United States), and from the centre of expertise in Belgium on alcohol and substance use (i.e., Flemish Expertise Centre Alcohol and Other Drugs). These three approaches resulted in a preliminary set of variables, which was the starting point for the expert consultation.

In a next step, an interdisciplinary expert panel was invited to contextualise the preliminary set of variables to the Belgian setting by providing input with regard, but not limited, to the definition of the variables, response options and formulations, and timing of the questions to be included in the BELpREG questionnaires. The panel consisted of seven Belgian experts in the field of obstetrics, neonatology/paediatrics (N = 2), midwifery, family medicine, perinatal psychiatry, and pharmacy, and were consulted in two rounds. In the first round, the experts completed an online survey (in Qualtrics; Augustus-September 2021) where they could indicate to which extent they agreed with any variables and/or response options; additional suggestions could be provided via open text fields. For each variable, experts were also asked to suggest the optimal timing and frequency of querying this variable. In the second round, the survey results were discussed in group during a real-life meeting, moderated by M.C. and L.S., to decide on inconsistent responses and/or any suggestions raised (September 2021).

### 2.3 Development of the BELpREG Infrastructure

To develop the BELpREG registration system, including the technical infrastructure and legal, privacy and ethical framework for data collection and management, collaboration with external partners was set-up. Legal consultants with expertise in data protection and IP law in the field of life sciences were involved, as well as the Data Protection Officer of our institute, solving questions related to the collection of health data compliant with applicable privacy legislations, database protection and the use of the collected data for research purposes. Besides, IT consultants with extensive expertise in data systems related to medicines contributed to the construction of the BELpREG registration system and facilitated the longitudinal framework for data collection and linkage to existing databases (in collaboration with the IT department within our institute, KU Leuven).

## 3 Results

### 3.1 BELpREG Data Categories and Variables

Overall, BELpREG encompasses questions on data variables categorized in one of the following seven categories: 1) Sociodemographic characteristics; 2) Information on the current pregnancy and health status; 3) Maternal-obstetric history; 4) Use of medicines, folic acid / pregnancy vitamins and other health products; 5) Substance use; 6) Pregnancy outcomes; and 7) Neonatal outcomes (see Table 1). Information on these categories will be longitudinally collected using questionnaires at different time points during and after pregnancy. A detailed overview of the complete list of BELpREG variables, per category, can be found in the Supplementary Material 1. To optimally understand the remainder of this manuscript, some basic information about the categories is provided in the following paragraphs.

**Table 1.**
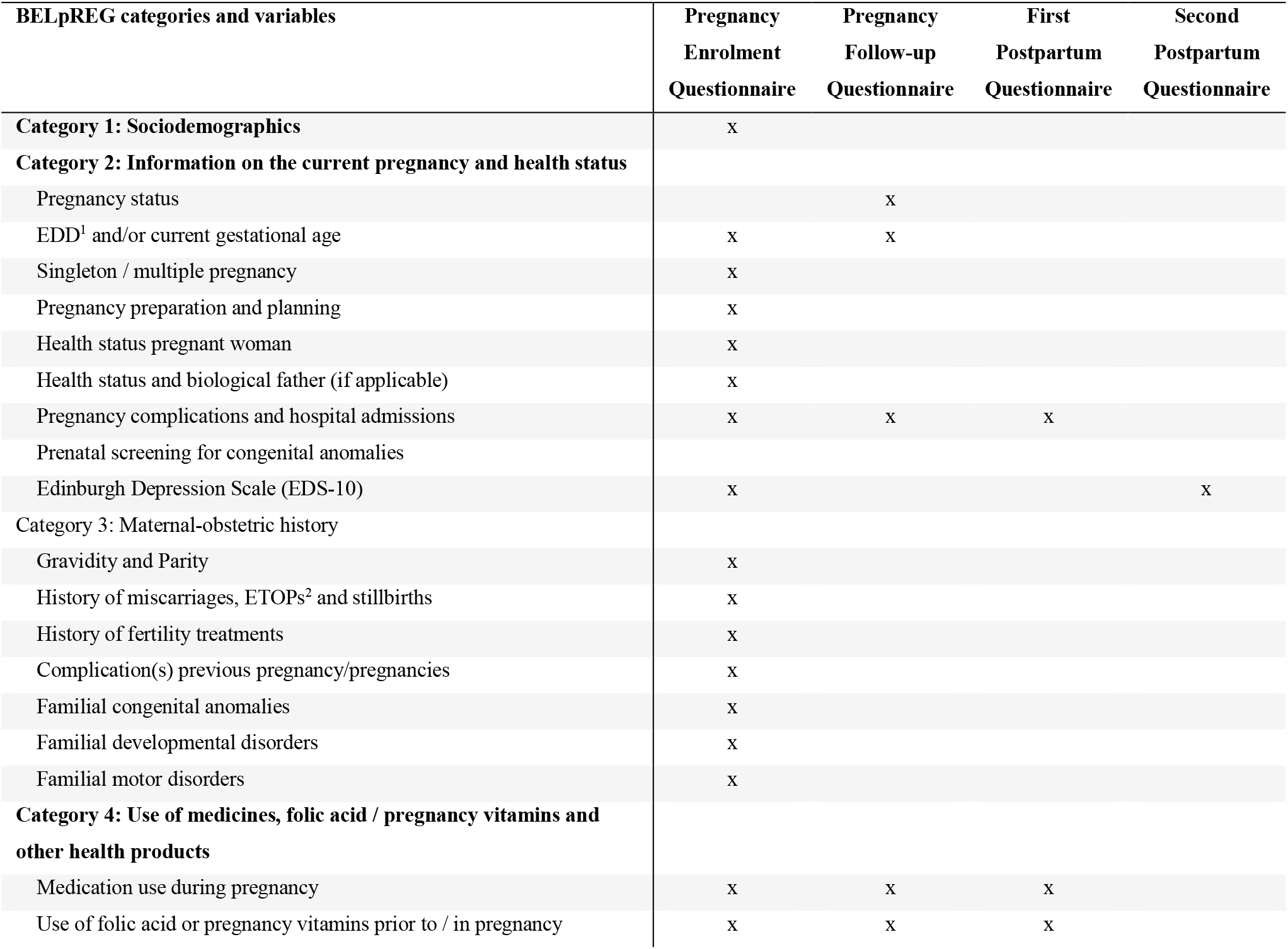
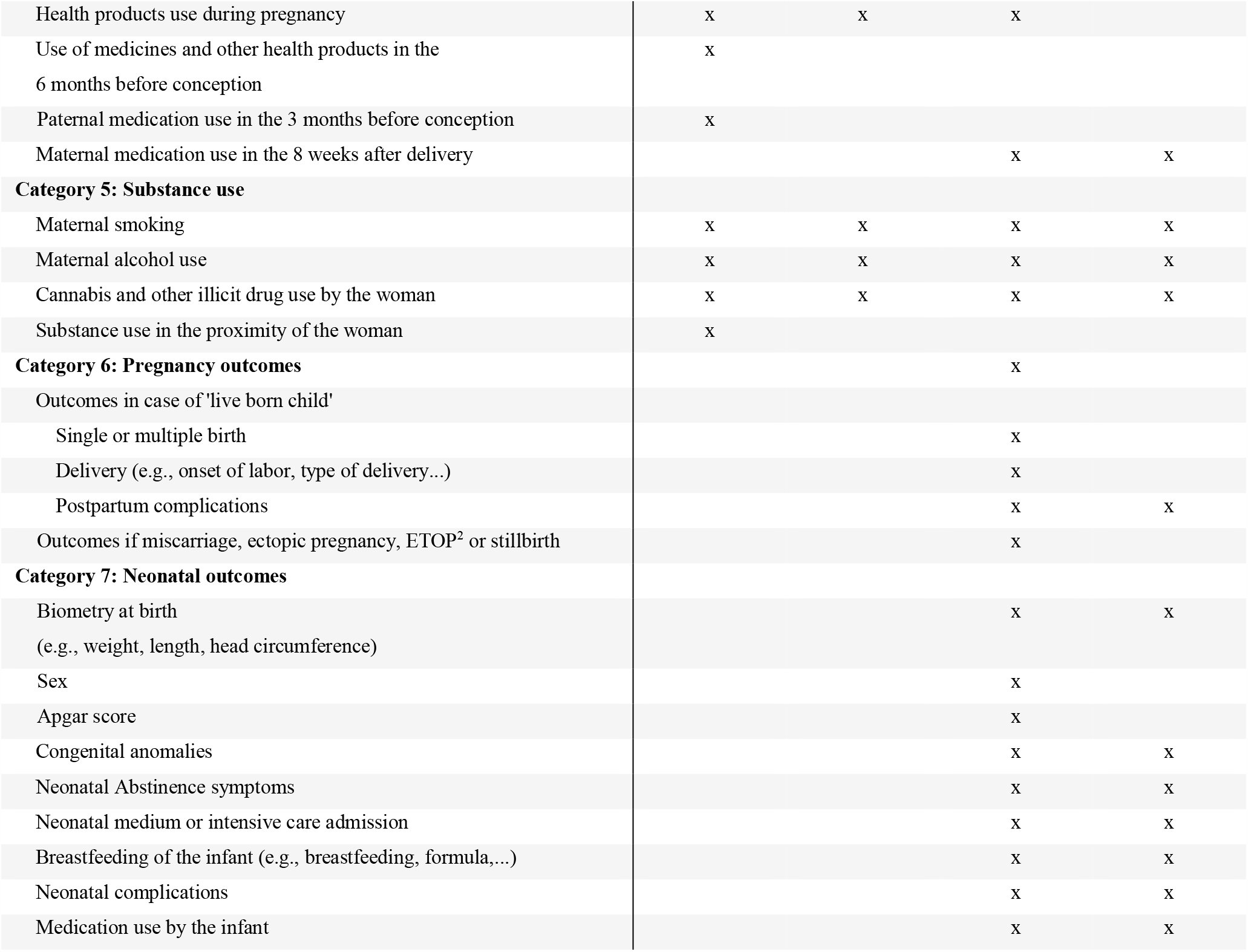
Overview of the overarching BELpREG categories and variables, according to their occurrence in one of the four types of survey instruments

1. The category *Sociodemographic characteristics* consists of basic identifiers of BELpREG participants, as for example, zip code, nationality, ethnicity, or the socio-economic status of the mother and biological father (the latter only if known).
2. *Information on the current pregnancy and health status* will provide details on the estimated date of delivery (EDD), gestational age at completion of the pregnancy follow-up questionnaires, plurality, pregnancy preparation/planning/follow-up, current and previous health status of mother and biological father (e.g., comorbidities), and pregnancy complications diagnosed so far. Further, this category includes a screening of depressive symptoms during pregnancy and the postnatal period with the Edinburgh Depression Scale (EDS) (Cox et al., 1987; Bergink et al., 2011). The EDS is considered the gold standard to detect PND (Cox et al., 1987). This scale was initially developed for the postpartum period but has also been validated for assessment during pregnancy (Bergink et al., 2011). It consists of a 10-item list regarding the severity of the depressive symptoms during the past 7 days.
3. The category *Maternal-obstetric history* includes details on previous pregnancies, including miscarriages, elective terminations of pregnancy (ETOP), stillbirths, complications in previous pregnancies and familial predisposition to congenital anomalies, developmental disorders, and motor disorders.
4. The category *Use of medicines, folic acid / pregnancy vitamins and other health products* is divided into the use of medication, folic acid and/or pregnancy vitamins, and other health products, including details on administration route, amount of the exposure (‘dose’) and exact timing of initiation and duration of exposure during pregnancy. Exposure to medicines and health products in the 6 months before the start of the current pregnancy is also questioned, along with paternal medication use in the 3 months before conception and maternal medication use in the first eight weeks postpartum.
5. The category *Substance use* includes variables questioning maternal exposure to alcohol, tobacco (i.e., smoking), cannabis, and other illicit substances. Besides, substance use in the proximity of the woman (‘passive exposure’) is covered as well, for example, by questioning about ‘in-house’ smoking by persons living together with the mother.
6. In the category *Pregnancy outcomes*, the addressed variables vary depending on the pregnancy outcome of the current pregnancy, i.e., the occurrence of a live birth, miscarriage, ectopic pregnancy, elective termination of pregnancy or stillbirth. In case of a live birth, additional questions about the delivery and potential postpartum complications are included.
7. The category *Neonatal outcomes* covers the health parameters of the infant, including the identification of congenital anomalies and the occurrence of neonatal complications, but also the feeding of the infant.

### 3.2 Longitudinal Framework for Data Collection

Determining the interval of successive questionnaires and the timing and frequency of addressing specific variables were key elements in the design of the BELpREG longitudinal framework. Following careful consideration, an interval of 4 weeks between two successive questionnaires was chosen, with data collection until 8 weeks postpartum, except in case of consent withdrawal or early pregnancy interruption.

Four different survey instruments, each with specific data variables, were constructed: i.e., A) the Pregnancy Enrolment Questionnaire; B) the Pregnancy Follow-up Questionnaire; C) the First Postpartum Questionnaire; and D) the Second Postpartum Questionnaire. Table 1 gives an overview of the categories and variables questioned in each of the different questionnaires.

Figure 2 provides a general overview of the BELpREG longitudinal framework, i.e., from enrolment to study execution and termination of participation. More practical details about the framework are provided in the following paragraph.

**Figure 2.**
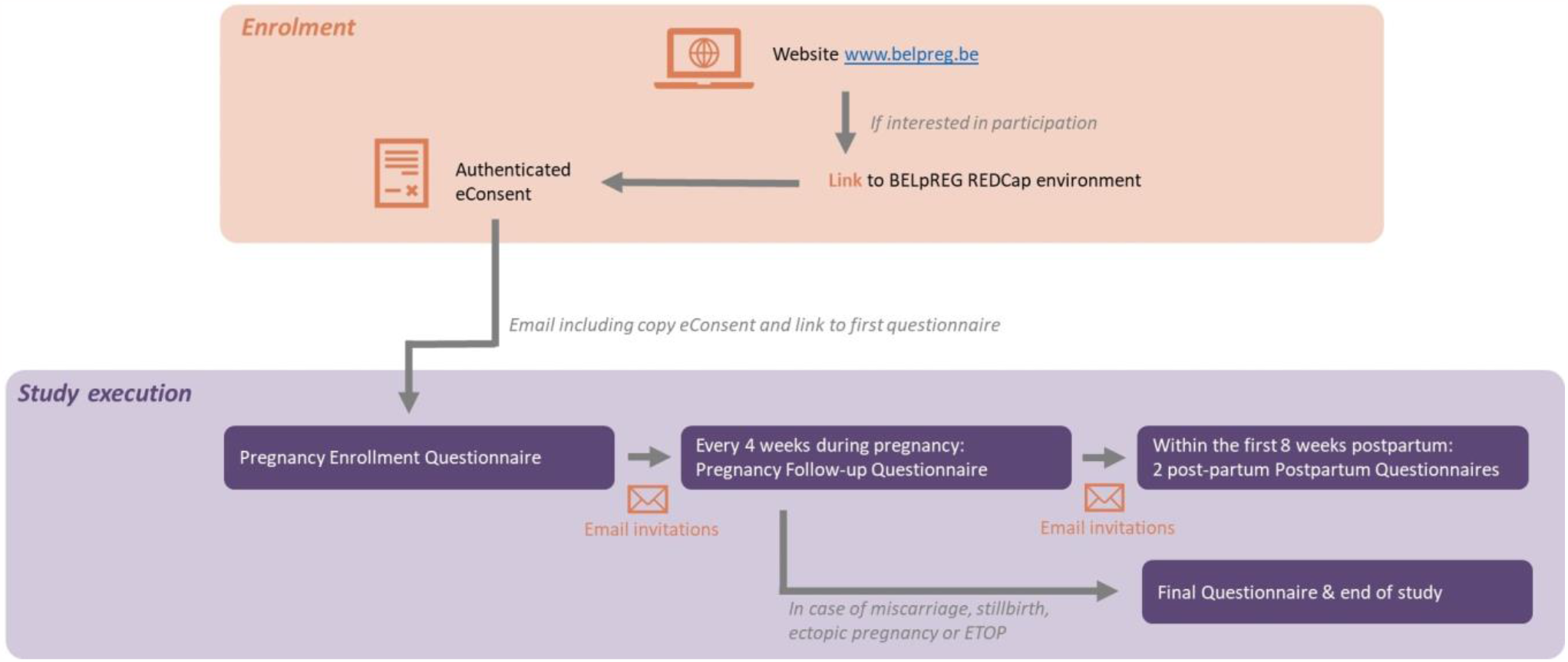
Overview of the BELpREG longitudinal framework, from enrolment to study execution

To start data-collection, self-enrolment by pregnant persons is required, by using the link on the BELpREG website (www.belpreg.be) or scanning the QR code on BELpREG flyers. Enrolment is possible at any time during pregnancy, but participants will be encouraged to start as soon as possible in pregnancy. Once a pregnant person has enrolled, a new record ID is automatically generated in the BELpREG database, allowing longitudinal data collection on individual level. Upon completion of the digital consent procedure (see 3.3.1) and appropriateness check of the eligibility criteria (i.e., being pregnant, ≥18 years and receiving healthcare in Belgium), the Pregnancy Enrolment Questionnaire immediately starts. This is the most comprehensive questionnaire to be completed. It may take 20-30 minutes or even more, depending on gestational age at enrolment, gravidity and the number of medicines used during the current pregnancy so far. Completion of the *Sociodemographic characteristics* in the Pregnancy Enrolment Questionnaire triggers the scheduling of the subsequent e-mail invitations for the Pregnancy Follow-up Questionnaires. These Pregnancy Follow-up Questionnaires are generally expected to require less time to be completed, i.e., ± 5-10 minutes, depending on the number of medicines, vitamins and health products used. The Pregnancy Follow-up Questionnaires mainly focus on medication and health products’ use since the latest completed questionnaire (i.e., ideally, four weeks before), but also contain questions on pregnancy complications and maternal substance use (see Table 1 1). If a Pregnancy Follow-up Questionnaire is not completed after two weeks, a reminder is automatically sent via e-mail. The invitations for subsequent Pregnancy Follow-up Questionnaires are sent out, irrespective of whether the previous questionnaire(s) were completed, but with a maximum of 10 invitations per person. Once a participant indicates that the pregnancy has ended, the First Postpartum Questionnaire immediately appears. In case a miscarriage, stillbirth, ectopic pregnancy, or pregnancy termination is registered, participants do not receive additional questionnaires for this pregnancy anymore. They can register themselves again in the event of a new pregnancy. Only if a live birth is registered in the First Postpartum Questionnaire, the Second Postpartum Questionnaire is sent out four weeks later. In case of a live born twins, the questions related to Neonatal outcomes are automatically duplicated in both Postpartum Questionnaires. A Second Postpartum Questionnaire is only sent out if the First Postpartum Questionnaire was completed in the first 28 days after birth.

Finally, the data entry forms in the category *Use of medicines, folic acid / pregnancy vitamins and other health products* have a longitudinal ‘memory function’. This means that, if daily used medicines or health products were registered, the names of these products as well as details related to its use are automatically shown in the next questionnaire(s). This functionality gives participants the opportunity to quickly and easily indicate if they still use the product in a similar way or not, and hence, to save time because of this easy registration.

### 3.3 Infrastructure of the BELpREG Registration System

The BELpREG registration system was built using Research Electronic Data Capture (REDCap) software (Harris et al., 2009; Harris et al., 2019; Van Bulck et al., 2022). REDCap is a secure, web-based, software platform, used to collect, manage, and store longitudinal research data. The BELpREG REDCap system, database and back-up system are hosted on an institutional server separated from other institutional REDCap projects, providing an additional layer of security. The BELpREG REDCap system adheres to high-level data security standards and requires a multi-factor authentication procedure for investigators to get access. Only pre-authorized users with fixed roles (*users rights*) have access to the BELpREG REDCap management tool and database. Access to REDCap records is automatically logged, and changes to recorded data are stored in *independent audit trails*. In BELpREG, REDCap’s security measures for identifiable data apply, i.e., *identifier tags* are attached to variables collecting identifiers, ensuring that only pseudonymized data will be extracted out of the database.

To fit the REDCap software to the specific needs of the BELpREG project, the following features have been created: 1) digital consent procedure (*eConsent*) (see 3.3.1), 2) linkage of BELpREG survey instruments with databases of medications and health products (see 3.3.2); and 3) linkage of BELpREG survey instruments with external databases (see 3.3.3), to facilitate both data entry, coding, and analysis, and to enhance user satisfaction.

#### 3.3.1 eConsent Procedure

As personal and health data are processed in the BELpREG project, the General Data Protection Regulation (GDPR) applies for each processing activity. Given the sensitive nature of the collected data, along with our ambitions with respect to data pooling as part of (international) collaborative projects, obtaining informed consent from participants prior to enrolment was necessary. However, as pregnant persons should be able to enrol without physical/in-person contact with the BELpREG researchers, the availability of a digital, ‘remote’ consent (‘eConsent’) was vital. Besides, according to applicable legislation in Belgium on research involving human subjects, authentication of participants was needed as part of the BELpREG consent procedure and required a simple solution.

The following eConsent procedure was developed using REDCap software and is currently in place (see Supplementary Material 2). First, potential participants watch an instructive video, available on the BELpREG website (www.belpreg.be), providing information on the objectives and course of the project, inclusion criteria, risks and benefits, consent withdrawal and confidential treatment of the collected data; this replaces the ‘in-person’ provision of study information as part of a regular informed consent process. In case of questions, the researchers can easily be contacted via the website, email, or phone. After watching the video, participants are redirected to the REDCap environment. Once individuals confirm that they watched the video and want to sign the consent form, an automatic e-mail is sent out containing a One-Time-Password (OTP), which should be used, as authentication procedure, to sign the form ‘remotely’. Once it is completed, a copy of the form is stored by the researchers and sent to the participant, who can immediately start with the Pregnancy Enrolment Questionnaire. This eConsent procedure, and the overall BELpREG research project, have been approved by the local Ethics Committee Research UZ/KU Leuven (S66464).

#### 3.3.2 Linked Databases of Medicines and Health Products

The survey instrument on *Use of medicines, folic acid / pregnancy vitamins and other health products* includes linkages with both the official Belgian medication database (i.e., “Source Authentique des Médicaments” or SAM) and the Medipim database (i.e., a database containing recent pictures of the packages of commercially available medicines and health products in Belgium). When participants enter the name of a medicine, vitamin, or other health product in a BELpREG questionnaire field, a structured and relevant dropdown list (based on both databases) appears that facilitates the selection of the right product. The linked databases allow data collection on medication, vitamin, and health products’ use in a structured, reliable, and user-friendly way. At the same time, by using underlying structured data fields, the technical details of the registered medicines and products, such as, for example, the Anatomical Therapeutic Chemical classification (ATC) code and the route of administration, are automatically extracted from the linked databases and imported into the BELpREG database.

#### 3.3.3 Other Linked Databases

In addition to the structured data entry of medicines, vitamins, and health products, other BELpREG variables also have underlying databases generating structured dropdown lists in their response field. This is the case for the registration of 1) country of residence and nationality (i.e., linked to the NATO database); 2) city of residence (i.e., linked to the database of the national post service in Belgium); 3) name of the hospital where the delivery took place (i.e., linked to the database of the National Institute for Health and Disability Insurance); and 4) the names of individual HCPs following-up BELpREG participants during pregnancy and in the postpartum period (i.e., also linked to a database of the latter national institute). Furthermore, to register (chronic) disorders or indications of medications, participants can choose from a list of frequently reported indications/disorders. This list was developed based on literature and clinical experience with pregnant women and will grow over time (with up to five updates per year) based on experiences obtained by the registrations of BELpREG participants using this list. In the back-end of the system, the indications/disorders are linked to their respective ICD-11 (International Classification of Diseases 11th Revision, n.d.) and MedDRA code (Medical Dictionary for Regulatory Activities, n.d.), facilitating data processing, analysis, and reporting, and data pooling.

## 4 Discussion

### 4.1 Development and Design of the BELpREG Registration System

In this paper, we described the development and design of the BELpREG registration system, enabling the prospective collection of ‘real-world’, observational data on perinatal medication use and mother-infant outcomes in Belgium. After almost two years of constructing the system, we succeeded in delivering the BELpREG infrastructure, including data variables, longitudinal framework, digital consent and linkage with medication and other databases. Given the current lack of routine data collection and pharmacoepidemiologic research on perinatal medication use in Belgium, BELpREG can be considered a unique registration system/initiative that merits attention from researchers, MAHs and policy makers in Belgium and beyond.

In the next paragraphs, we will discuss in detail the value of the BELpREG variables (see 4.1.1), longitudinal framework and infrastructure (see 4.1.2) and recruitment of pregnant persons (see 4.1.3) and will elaborate on potential strengths and limitations of the system.

#### 4.1.1 The BELpREG Variables

The current tools used for the registration of medication exposure in pregnancy and mother-infant outcomes, as part of international research initiatives on perinatal medication safety, often differ in terms of exposure and outcome variables, including clinical definitions and coding standards (Thurin et al., 2022). Such registration variety clearly complicates data pooling across countries, necessitating additional standardization and alignment procedures with the inherent risk of considerable loss of data and/or granularity. Data pooling is, however, very important in this field to enlarge the sample size and power to detect significant associations, especially when studying rare outcomes such as (specific types of) congenital malformations (Gelperin et al., 2017).

Relying on relevant documents on medication safety in pregnancy research, such as the IMI ConcePTION Core Data Elements (ConcePTION, n.d.), we pursued and eventually delivered a list of variables that are in line with those applied in other international data collection initiatives, facilitating data pooling in the future. Contextualization to the Belgian setting was, however, considered appropriate, to maximize the feasibility and likelihood of obtaining complete and reliable data for all variables and to ensure user involvement and satisfaction. Contextualization was done with input from an interdisciplinary panel consisting of experts from the main professions involved in perinatal care. Despite the large number of variables, which may be cumbersome for participants, the integration of a broad set of potential confounders for associations between medication use and pregnancy outcomes is a strength of the BELpREG system. In fact, observational studies using health utilisation data typically lack information on many confounders, entailing limitations in terms of the validity of the conclusions (Huybrechts et al., 2019; Panchaud et al., 2022). With respect to the feeding status of the infant, breastfeeding or receiving breast milk will be asked, which is considered an important but often ignored source of medication exposure, a mitigator of adverse effects or a relevant covariate in pharmacoepidemiologic research on the long-term effects of medication use in pregnancy (Jordan et al., 2022).

#### 4.1.2 The BELpREG Longitudinal Framework and Technical Infrastructure

As the ambition is to use BELpREG data for longitudinal research, the BELpREG system needed to consist of online questionnaires which are sent and (ideally) completed by pregnant persons at multiple time points during and after pregnancy. We opted for a fixed interval of four weeks, lasting – at least for now – until eight weeks postpartum. By choosing this time interval, we tried to find the right balance between registration fatigue among participants and recall bias for the self-reported registration of medication and/or health products used in the previous period (van Gelder et al., 2018). Upon enrolment, BELpREG participants are prospectively followed-up, i.e., they register exposure data throughout pregnancy without knowing the pregnancy outcome yet. Collecting data in a prospective way is an important strength in observational research, potentially decreasing the risk of bias (Schaefer et al., 2008). Yet, in the Pregnancy Enrolment Questionnaire, we ask what medicines/health products were used from the start of pregnancy until then. In case of late enrolment during pregnancy (i.e., at more advanced gestational age), the risk of recall bias could be very relevant. In any case, in all our communication, we advise and encourage individuals to enrol as early as possible during pregnancy, although enrolment is possible at any gestational age.

To support participants in registering their medicines and health products, a linkage with databases of medicines and health products available in Belgium was built, including pictures of the drug packages (see 3.3.2). At user level, these linkages may improve the user friendliness of the system (i.e., by facilitating the registration of their products from a dropdown list). At system level, the linkages with medication and other types of databases, for example related to nationality or zip-code, may have positive effects in terms of correctness/quality of the collected data (i.e., by selecting/recognizing the correct product by means of its picture) and ease of further data processing (i.e., by automatically importing relevant parameters, such as ATC code, in the BELpREG database).

The retained variables, linked databases and longitudinal framework were built into REDCap software, which is an internationally recognized research tool for data collection and management (Harris et al., 2009; Harris et al., 2019; Van Bulck et al., 2022), facilitating data collection initiatives in other settings or countries in the future by relying on the BELpREG infrastructure.

When defining the BELpREG design, the perspectives of MAHs towards medication safety monitoring and pharmacovigilance during pregnancy have been considered (Sillis et al., 2022), thereby maximizing the added value of BELpREG for MAHs and facilitating future collaborations by means of contract research with, for example, the aim to update the pregnancy statements in the product labels (Roque Pereira et al., 2022).

Nevertheless, when using the BELpREG data for descriptive/analytical pharmacoepidemiologic purposes, it should be noted that the reliability of the conclusions depends on the quality of the self-reported data. Hence, assessing the correctness of the collected data by means of a validation study is warranted and has already been initiated (see 4.2).

#### 4.1.3 Recruitment of Pregnant Individuals

The success of ‘citizen science’ initiatives such as BELpREG depends on the level of involvement by pregnant participants. Likewise, its success also lies in collaborating with other data collection initiatives abroad, especially as Belgium is a small country with ‘only’ 120 000 pregnant women per year (Statbel, 2023). Therefore, difficulties can be expected to obtain enough exposed women, solely based on data collection in our country, to draw firm conclusions on associations. Data pooling will be key, also for timely signal detection. Nevertheless, capturing every single piece of data is ethically appropriate and must be pursued.

To reach pregnant persons, also those who are not using (chronic) medicines, applying diverse recruitment methods is vital (Goldstein et al., 2021). Therefore, to date, pregnant persons are informed about BELpREG both in-person (direct, by HCPs) and online (indirect). First, details about BELpREG are being shared with different types of HCPs involved in perinatal care (i.e., obstetricians, midwives, general practitioners, pharmacists…), with support from their professional organizations and regional care organizations. The embeddedness of BELpREG in the local healthcare context for pregnant persons may be a critical aspect for its successful uptake by potential users (Sillis et al., 2022). Sustainable partnerships between the research team and HCPs, prenatal care organizations and hospitals has earlier been shown as a core strategy for successful recruitment (Goldstein et al., 2021), and are currently being set-up in within BELpREG. Second, indirect recruitment strategies are being used by means of our website, social media (Facebook, Instagram), and printed/digital posters and flyers. As poor public awareness of research among different groups within the population is a large barrier for participation in research (Goldstein et al., 2021), social media can be an effective method to reach a broader audience, independent from efforts by HCPs. In fact, the population of pregnant women often uses social media to search for information (Sinclair et al., 2018). In addition, high internet penetration rates exist among women of childbearing age in Belgium (Statbel, 2022). Previous research has shown the advantages of recruiting pregnant women through social media, including advertisement via these channels (van Gelder et al., 2019). Although social media seem a logical recruitment channel for BELpREG, its effectiveness should be further explored, including the effect of directly approaching the target population via (advertising using) social media (Sarker et al., 2017).

As study enrolment in BELpREG occurs without physical contact with the researchers, the ability to provide remote informed consent is paramount. Therefore, in BELpREG, a user-friendly and GDPR compliant eConsent framework was specifically developed according to the applicable legislation in Belgium (see 3.3.1). The use of an eConsent in REDCap has previously been shown to be feasible and easy-to-use by pregnant women (Phillippi et al., 2018). In the future, the BELpREG eConsent framework could serve as an instrument or starting point for similar research purposes in other populations and/or settings.

### 4.2 Future Perspectives of BELpREG

As part of the development stage, the BELpREG infrastructure was extensively tested ‘in-house’ on functionality and technical performance by different persons and by using different scenarios. In November 2022, public data collection has officially started. However, as the next step, we have planned to closely follow-up and explore the ongoing data collection in ‘real-life’ and the preliminary data. Such iterative process of testing, followed by possible system modifications, will result in a robust, patient-friendly, and fit-for-purpose research instrument, ready for large-scale implementation in Belgium and beyond. The assessment will focus on 1) the completeness of the collected data, both within (i.e., missing variables) and across questionnaires (i.e., loss to follow-up); 2) the ‘reach’ or representativeness of the BELpREG sample (as research using online questionnaires typically only reaches a select group of the population (Vorstenbosch et al., 2019; van Gelder et al., 2020; Stegherr et al., 2022)); 3) users’ experiences and challenges with data entry (by means of a mixed-methods approach); 4) experiences of HCPs with informing and motivating pregnant individuals to participate in BELpREG.

In addition to the feasibility assessment, we will undertake a validation study to evaluate the quality of the self-reported data. Patients may probably be best placed to record their medication use (van Gelder et al., 2018), however, it remains unclear to what extent they are able to correctly register pregnancy and neonatal outcomes (e.g., type and description of congenital anomalies). Therefore, the validity of self-registered health data will be assessed by comparing with the data available in medical/obstetric records (only for participants who gave consent to contact their HCPs). In case the validation study shows that these variables can indeed be reliably registered by patients, this will avoid an additional registration burden for HCPs. Overall, it should be further explored to what extent HCPs would be able to register data in BELpREG themselves, for example for patients with low health literacy or impaired internet access.

Another important future perspective relates to the exploration for long-term follow-up of children born after prenatal exposure to medicines. To date, the BELpREG design only includes maximum two questionnaires in the first eight weeks postpartum, addressing neonatal health outcomes. However, some exposures may result in adverse neurodevelopmental outcomes in offspring, which only become visible months or years after birth (Liu et al., 2017; Bromley and Bluett-Duncan, 2021). We have the ambition to extend our postpartum questionnaires for long-term follow-up purposes, and therefore look expectantly at the ongoing LIFETIME project (i.e., Long-term Investigation Following Exposure To Individual Medicines in utEro) (ConcePTION, n.d.). As a first step, BELpREG participants are being asked about their willingness to complete self-reported surveys at the age of developmental milestones (Hjorth et al., 2019).

Finally, translating the BELpREG questionnaires into other (national) languages (e.g., French, English) is also an intended goal, which we hope to achieve in the (near) future.

## 5 Conclusions

Due to the current lack of routine data collection and pharmacoepidemiologic research on medication safety in pregnancy in Belgium, the BELpREG registration system was developed. As a ‘citizen science’ project, the BELpREG system enables the prospective and longitudinal collection of comprehensive, real-world data on perinatal medication use and mother-infant outcomes. Online questionnaires will be completed by pregnant persons every four weeks during pregnancy and in the first eight weeks postpartum.

The variables were compiled using relevant documents and through an expert panel consultation and are structured in seven categories. A digital, ‘remote’ informed consent and linkage to medication databases, with images of drug packages and underlying structured data fields, are built into the system, which uses REDCap as software. Data collection has officially started in November 2022.

Based on its rigorous and fully digital design, BELpREG holds the potential to be a successful, sustainable, and collaborative research tool, enabling perinatal pharmacoepidemiologic research in Belgium and beyond. Researchers, MAHs and policy makers are invited to contact the researchers to explore future collaborations and data pooling, with the aim to reduce the current knowledge gap in this field and to provide more evidence on medication safety in pregnancy to patients and HCPs.

## Data Availability

Not applicable.

## 6 Conflict of Interest

The authors declare that the research was conducted in the absence of any commercial or financial relationships that could be construed as a potential conflict of interest.

## 7 Supplementary Material

The following supporting information can be downloaded: Supplementary Table1: Detailed overview of the BELpREG variables, per category; Supplementary Figure 2: Schematic overview of the eConsent procedure.

## 8 Author Contributions

Conceptualization, L.S., V.F. and M.C.; methodology, L.S., V.F., K.A., A.B., M.D.V., T.H., A.S., K.V.C., J.Y.V. and M.C.; software, L.S. and M.C.; validation, L.S., V.F. and M.C.; formal analysis, L.S. and M.C.; investigation, L.S. and M.C.; resources, V.F., M.C. and L.S.; data curation, L.S.; writing-original draft preparation, L.S. and M.C.; writing-review and editing, V.F., K.A., A.B., M.D.V., T.H., A.S., K.V.C., and J.Y.V.; supervision, V.F. and M.C.; project administration, L.S. and M.C.; funding acquisition, V.F., M.C. and L.S.. All authors have read and agreed to the published version of the manuscript.

## 9 Funding

The development of BELpREG was funded by a KU Leuven internal fund (C3/020/095). The research activities of Laure Sillis are funded by the Research Foundation Flanders (FWO) (1S35823N). The research activities of Michael Ceulemans are supported by the Department of Pharmaceutical and Pharmacological Sciences and the Faculty of Pharmaceutical Sciences of the KU Leuven.

## 10 Acknowledgments

The authors would like to thank the following persons for their input and help through the development of BELpREG: Hilde De Tollenaere and Peter Van Rompaey (Clinical Trial Center, University Hospitals Leuven), Eugène Van Puijenbroek (Lareb, NL), Saskia Vorstenbosch (NL), Carlijn Litjens (NL), Ellen Ederveen (Lareb, NL), Judith Hendriks (Lareb, NL), Jonathan Luke Richardson (UKTIS, UK), Rebecca Bromley and Matthew Bluett-Duncan (University of Manchester, UK), Liesbet Van Bulck (KU Leuven), Alexis Puvrez (KU Leuven) and Gert Goos (ICT Service, KU Leuven). The authors are also grateful to the BELpREG partners Digile (IT support - An Crepel, Karel De Loof and Davy Hollevoet), Medipim (providing pictures of medicinal products), Allen&Overy (legal advice) and Isabelle Geeraerts (graphical design). Finally, the authors thank all colleagues and women who have contributed to the in-house testing of the system.

## 12 Data Availability Statement

Not applicable.

## 11 Note from the authors

The list of used REDCap features is not exhaustive. The mentioned features of REDCap used in the BELpREG project are a choice of the authors. Complete information about REDCap features can be found here: https://projectredcap.org/.

## Notes

### Competing Interest Statement

The authors have declared no competing interest.

### Author Declarations

This eConsent procedure, and the overall BELpREG research project, have been approved by the local Ethics Committee Research UZ/KU Leuven (S66464).

## References

Adam, M.P., Polifka, J.E., and Friedman, J.M. (2011). Evolving knowledge of the teratogenicity of medications in human pregnancy. American Journal of Medical Genetics Part C: Seminars in Medical Genetics 157(3), 175–182. doi: 10.1002/ajmg.c.30313.

Benevent, J., Hurault-Delarue, C., Araujo, M., Montastruc, J.L., Lacroix, I., and Damase-Michel, C. (2019). POMME: The New Cohort to Evaluate Long-Term Effects After Prenatal Medicine Exposure. Drug Saf 42(1), 45–54. doi: 10.1007/s40264-018-0712-9.

Bergink, V., Kooistra, L., Lambregtse-van den Berg, M.P., Wijnen, H., Bunevicius, R., van Baar, A., et al. (2011). Validation of the Edinburgh Depression Scale during pregnancy. J Psychosom Res 70(4), 385–389. doi: 10.1016/j.jpsychores.2010.07.008.

Bromley, R.L., and Bluett-Duncan, M. (2021). Neurodevelopment Following Exposure to Antiseizure Medications in Utero: A Review. Curr Neuropharmacol 19(11), 1825–1834. doi: 10.2174/1570159x19666210716111814.

Ceulemans, M., Fortuin, M., Van Calsteren, K., Allegaert, K., and Foulon, V. (2020). Prevalence and characteristics of pregnancy-and lactation-related calls to the National Poison Centre in Belgium: A retrospective analysis of calls from 2012 to 2017. Journal of Evaluation in Clinical Practice 26(3), 911–917. doi: 10.1111/jep.13228.

Ceulemans, M., Foulon, V., Panchaud, A., Winterfeld, U., Pomar, L., Lambelet, V., et al. (2022a). Self-Reported Medication Use among Pregnant and Breastfeeding Women during the COVID-19 Pandemic: A Cross-Sectional Study in Five European Countries. International Journal of Environmental Research and Public Health 19(3), 1389. doi: 10.3390/ijerph19031389.

Ceulemans, M., Van Calsteren, K., Allegaert, K., and Foulon, V. (2022b). Information Needs and Counseling Preferences among Potential Users of the Future Teratology Information Service in Belgium: A Cross-Sectional Study Involving the Public and Healthcare Professionals. International Journal of Environmental Research and Public Health 19(14), 8605. doi: 10.3390/ijerph19148605.

Chambers, C.D., Johnson, D.L., Xu, R., Luo, Y., Lopez-Jimenez, J., Adam, M.P., et al. (2019). Birth outcomes in women who have taken adalimumab in pregnancy: A prospective cohort study. PLOS ONE 14(10), e0223603. doi: 10.1371/journal.pone.0223603.

ConcePTION. (n.d.). https://www.imi-conception.eu/ [Accessed February 14, 2023].

Cox, J.L., Holden, J.M., and Sagovsky, R. (1987). Detection of Postnatal Depression. British Journal of Psychiatry 150(6), 782–786. doi: 10.1192/bjp.150.6.782.

Dandjinou, M., Sheehy, O., and Bérard, A. (2019). Antidepressant use during pregnancy and the risk of gestational diabetes mellitus: a nested case–control study. BMJ Open 9(9), e025908. doi: 10.1136/bmjopen-2018-025908.

Dao, K., Shechtman, S., Diav-Citrin, O., George, N., Richardson, J.L., Rollason, V., et al. (2022). Reproductive Safety of Trazodone After Maternal Exposure in Early Pregnancy: A Comparative ENTIS Cohort Study. Journal of Clinical Psychopharmacology, 10.1097/JCP.0000000000001630. doi: 10.1097/jcp.0000000000001630.

EMA (2019). “GVP Product-or Population-Specific Considerations III: Pregnant and breastfeeding women (EMA/653036/2019) - Draft for public consultation”.).

Gelperin, K., Hammad, H., Leishear, K., Bird, S.T., Taylor, L., Hampp, C., et al. (2017). A systematic review of pregnancy exposure registries: examination of protocol-specified pregnancy outcomes, target sample size, and comparator selection. Pharmacoepidemiology and Drug Safety 26(2), 208–214. doi: 10.1002/pds.4150.

Gerbier, E., Favre, G., Tauqeer, F., Winterfeld, U., Stojanov, M., Oliver, A., et al. (2022). Self-Reported Medication Use among Pregnant and Postpartum Women during the Third Wave of the COVID-19 Pandemic: A European Multinational Cross-Sectional Study. International Journal of Environmental Research and Public Health 19(9), 5335. doi: 10.3390/ijerph19095335.

Gerbier, E., Graber, S.M., Rauch, M., Marxer, C.A., Meier, C.R., Baud, D., et al. (2021). Use of drugs to treat symptoms and acute conditions during pregnancy in outpatient care in Switzerland between 2014 and 2018: analysis of Swiss healthcare claims data. Swiss Med Wkly 151, w30048. doi: 10.4414/smw.2021.w30048.

Goldstein, E., Bakhireva, L.N., Nervik, K., Hagen, S., Turnquist, A., Zgierska, A.E., et al. (2021). Recruitment and retention of pregnant women in prospective birth cohort studies: A scoping review and content analysis of the literature. Neurotoxicology and Teratology 85, 106974. doi: https://doi.org/10.1016/j.ntt.2021.106974.

Harris, P.A., Taylor, R., Minor, B.L., Elliott, V., Fernandez, M., O’Neal, L., et al. (2019). The REDCap consortium: Building an international community of software platform partners. Journal of Biomedical Informatics 95, 103208. doi: 10.1016/j.jbi.2019.103208.

Harris, P.A., Taylor, R., Thielke, R., Payne, J., Gonzalez, N., and Conde, J.G. (2009). Research electronic data capture (REDCap)—A metadata-driven methodology and workflow process for providing translational research informatics support. Journal of Biomedical Informatics 42(2), 377–381. doi: 10.1016/j.jbi.2008.08.010.

Hernandez-Diaz, S., and Oberg, A.S. (2015). Are Epidemiological Approaches Suitable to Study Risk/Preventive Factors for Human Birth Defects? Current Epidemiology Reports 2(1), 31–36. doi: 10.1007/s40471-015-0037-5.

Hernandez-Diaz, S., Smith, C.R., Shen, A., Mittendorf, R., Hauser, W.A., Yerby, M., et al. (2012). Comparative safety of antiepileptic drugs during pregnancy. Neurology 78(21), 1692–1699. doi: 10.1212/wnl.0b013e3182574f39.

Hjorth, S., Bromley, R., Ystrom, E., Lupattelli, A., Spigset, O., and Nordeng, H. (2019). Use and validity of child neurodevelopment outcome measures in studies on prenatal exposure to psychotropic and analgesic medications – A systematic review. PLOS ONE 14(7), e0219778. doi: 10.1371/journal.pone.0219778.

Huybrechts, K.F., Bateman, B.T., and Hernández-Dáz, S. (2019). Use of real-world evidence from healthcare utilization data to evaluate drug safety during pregnancy. Pharmacoepidemiology and Drug Safety 28(7), 906–922. doi: 10.1002/pds.4789.

Huybrechts, K.F., Bateman, B.T., Zhu, Y., Straub, L., Mogun, H., Kim, S.C., et al. (2021). Hydroxychloroquine early in pregnancy and risk of birth defects. American Journal of Obstetrics and Gynecology 224(3), 290.e291-290.e222. doi: 10.1016/j.ajog.2020.09.007.

International Classification of Diseases 11th Revision (ICD-11). (n.d.). https://icd.who.int/en [Accessed February 14, 2023].

Jordan, S., Bromley, R., Damase-Michel, C., Given, J., Komninou, S., Loane, M., et al. (2022). Breastfeeding, pregnancy, medicines, neurodevelopment, and population databases: the information desert. International Breastfeeding Journal 17(1). doi: 10.1186/s13006-022-00494-5.

Lacroix, I., Hurault, C., Sarramon, M.F., Guitard, C., Berrebi, A., Grau, M., et al. (2009). Prescription of drugs during pregnancy: a study using EFEMERIS, the new French database. European Journal of Clinical Pharmacology 65(8), 839–846. doi: 10.1007/s00228-009-0647-2.

Larcin, L., Lona, M., Karakaya, G., Van Espen, A., Damase-Michel, C., and Kirakoya-Samadoulougou, F. (2021). Using administrative healthcare database records to study trends in reimbursed medication dispensed during pregnancy in Belgium from 2003 to 2017. Pharmacoepidemiol Drug Saf. doi: 10.1002/pds.5299.

Liu, X., Agerbo, E., Ingstrup, K.G., Musliner, K., Meltzer-Brody, S., Bergink, V., et al. (2017). Antidepressant use during pregnancy and psychiatric disorders in offspring: Danish nationwide register based cohort study. BMJ, j3668. doi: 10.1136/bmj.j3668.

Lupattelli, A., Spigset, O., Twigg, M.J., Zagorodnikova, K., Mårdby, A.C., Moretti, M.E., et al. (2014). Medication use in pregnancy: a cross-sectional, multinational web-based study. BMJ Open 4(2), e004365. doi: 10.1136/bmjopen-2013-004365.

Magnus, P., Irgens, L.M., Haug, K., Nystad, W., Skjærven, R., and Stoltenberg, C. (2006). Cohort profile: The Norwegian Mother and Child Cohort Study (MoBa). International Journal of Epidemiology 35(5), 1146–1150. doi: 10.1093/ije/dyl170.

Medical Dictionary for Regulatory Activities (MedDRA). (n.d.). https://www.meddra.org/ [Accessed February 14, 2023].

Moseholm, E., Katzenstein, T.L., Pedersen, G., Johansen, I.S., Wienecke, L.S., Storgaard, M., et al. (2022). Use of antiretroviral therapy in pregnancy and association with birth outcome among women living with HIV in Denmark: A nationwide, population-based cohort study. HIV Medicine 23(9), 1007–1018. doi: 10.1111/hiv.13304.

Nörby, U., Noël-Cuppers, B., Hristoskova, S., Desai, M., Härmark, L., Steel, M., et al. (2021). Online information discrepancies regarding safety of medicine use during pregnancy and lactation: an IMI ConcePTION study. Expert Opinion on Drug Safety 20(9), 1117–1124. doi: 10.1080/14740338.2021.1935865.

Panchaud, A., Cleary, B., Weber-Schoendorfer, C., Shechtman, S., Cassina, M., Diav-Citrin, O., et al. (2022). The risk of questioning the safety of drugs considered safe in pregnancy at the era of big data: the everlasting case of doxylamine. J Clin Epidemiol 152, 125–126. doi: 10.1016/j.jclinepi.2022.10.007.

Pauliat, E., Onken, M., Weber-Schoendorfer, C., Rousson, V., Addor, M.-C., Baud, D., et al. (2021). Pregnancy outcome following first-trimester exposure to fingolimod: A collaborative ENTIS study. Multiple Sclerosis Journal 27(3), 475–478. doi: 10.1177/1352458520929628.

Phillippi, J.C., Doersam, J.K., Neal, J.L., and Roumie, C.L. (2018). Electronic Informed Consent to Facilitate Recruitment of Pregnant Women Into Research. Journal of Obstetric, Gynecologic & Neonatal Nursing 47(4), 529–534. doi: 10.1016/j.jogn.2018.04.134.

Roque Pereira, L., Durán, C.E., Layton, D., Poulentzas, G., Lalagkas, P.-N., Kontogiorgis, C., et al. (2022). A Landscape Analysis of Post-Marketing Studies Registered in the EU PAS Register and http://ClinicalTrials.gov Focusing on Pregnancy Outcomes or Breastfeeding Effects: A Contribution from the ConcePTION Project. Drug Safety 45(4), 333–344. doi: 10.1007/s40264-022-01154-7.

Sarker, A., Chandrashekar, P., Magge, A., Cai, H., Klein, A., and Gonzalez, G. (2017). Discovering Cohorts of Pregnant Women From Social Media for Safety Surveillance and Analysis. Journal of Medical Internet Research 19(10), e361. doi: 10.2196/jmir.8164.

Scaffidi, J., Mol, B., and Keelan, J. (2017). The pregnant women as a drug orphan: a global survey of registered clinical trials of pharmacological interventions in pregnancy. BJOG: An International Journal of Obstetrics & Gynaecology 124(1), 132–140. doi: 10.1111/1471-0528.14151.

Schaefer, C., Ornoy, A., Clementi, M., Meister, R., and Weber-Schoendorfer, C. (2008). Using observational cohort data for studying drug effects on pregnancy outcome--methodological considerations. Reprod Toxicol 26(1), 36–41. doi: 10.1016/j.reprotox.2008.05.064.

Shields, K.E., and Lyerly, A.D. (2013). Exclusion of pregnant women from industry-sponsored clinical trials. Obstet Gynecol 122(5), 1077–1081. doi: 10.1097/AOG.0b013e3182a9ca67.

Sillis, L., Foulon, V., Verbakel, J.Y., and Ceulemans, M. (2022). Experiences and Perspectives of Marketing Authorisation Holders towards Medication Safety Monitoring during Pregnancy: A Pan-European Qualitative Analysis. International Journal of Environmental Research and Public Health 19(7), 4248. doi: 10.3390/ijerph19074248.

Sinclair, M., Lagan, B.M., Dolk, H., and McCullough, J.E.M. (2018). An assessment of pregnant women’s knowledge and use of the Internet for medication safety information and purchase. Journal of Advanced Nursing 74(1), 137–147. doi: 10.1111/jan.13387.

Statbel. (2022). ICT use in households in Belgium. https://statbel.fgov.be/nl/themas/huishoudens/ict-gebruik-huishoudens [Accessed February 14, 2023].

Statbel. (2023). Births and Fertility in Belgium. https://statbel.fgov.be/nl/themas/bevolking/geboorten-en-vruchtbaarheid [Accessed February 14, 2023].

Stegherr, R., Beck, E., Hultzsch, S., Schaefer, C., and Dathe, K. (2022). Can non-responding mask or mimic drug effects on pregnancy outcome? Evaluation of case characteristics based on the national Embryotox cohort. Reproductive Toxicology. doi: 10.1016/j.reprotox.2022.05.013.

Thurin, N.H., Pajouheshnia, R., Roberto, G., Dodd, C., Hyeraci, G., Bartolini, C., et al. (2022). From Inception to ConcePTION: Genesis of a Network to Support Better Monitoring and Communication of Medication Safety During Pregnancy and Breastfeeding. Clinical Pharmacology & Therapeutics 111(1), 321–331. doi: 10.1002/cpt.2476.

Van Bulck, L., Wampers, M., and Moons, P. (2022). Research Electronic Data Capture (REDCap): tackling data collection, management, storage, and privacy challenges. European Journal of Cardiovascular Nursing 21(1), 85–91. doi: 10.1093/eurjcn/zvab104.

van Gelder, M.M.H.J., Merkus, P.J.F.M., Van Drongelen, J., Swarts, J.W., Van De Belt, T.H., and Roeleveld, N. (2020). The PRIDE Study: Evaluation of online methods of data collection. Paediatric and Perinatal Epidemiology 34(5), 484–494. doi: 10.1111/ppe.12618.

van Gelder, M.M.H.J., Rog, A., Bredie, S.J.H., Kievit, W., Nordeng, H., and Belt, T.H. (2019). Social media monitoring on the perceived safety of medication use during pregnancy: A case study from the Netherlands. British Journal of Clinical Pharmacology 85(11), 2580–2590. doi: 10.1111/bcp.14083.

van Gelder, M.M.H.J., Van De Belt, T.H., Engelen, L.J.L.P.G., Hooijer, R., Bredie, S.J.H., and Roeleveld, N. (2019). Google AdWords and Facebook Ads for Recruitment of Pregnant Women into a Prospective Cohort Study With Long-Term Follow-Up. Maternal and Child Health Journal 23(10), 1285–1291. doi: 10.1007/s10995-019-02797-2.

van Gelder, M.M.H.J., Vorstenbosch, S., Te Winkel, B., van Puijenbroek, E.P., and Roeleveld, N. (2018). Using Web-Based Questionnaires to Assess Medication Use During Pregnancy: A Validation Study in 2 Prospectively Enrolled Cohorts. Am J Epidemiol 187(2), 326–336. doi: 10.1093/aje/kwx239.

Vorstenbosch, S., Te Winkel, B., van Gelder, M.M.H.J., Kant, A., Roeleveld, N., and van Puijenbroek, E. (2019). Aim and Design of pREGnant, the Dutch Pregnancy Drug Register. Drug Saf 42(1), 1–12. doi: 10.1007/s40264-018-0722-7.

Zhao, Y., Du, G., Luan, X., Yang, H., Zhang, Q., Zhang, Z., et al. (2022). Registered Clinical Trials Comprising Pregnant Women in China: A Cross-Sectional Study. Frontiers in Pharmacology 13. doi: 10.3389/fphar.2022.850080.

