## Supplementary Table and Figure for "Development and design of the BELpREG registration system for the collection of real-world data on medication use in pregnancy and mother-infant outcomes"

### 1 Supplementary Table

Overview of the BELpREG data variables, shown per category and indicating in which survey instruments they are asked (i.e., time points at which the information is collected; EM = Pregnancy Enrolment Questionnaire; FU = Pregnancy Follow-up Questionnaire; PP 1 = First Postpartum Questionnaire; PP 2 = Second Postpartum Questionnaire). This overview follows, where possible, the order of the variables as they appear in the BELpREG questionnaires.

| **Category 1: Sociodemographics** | | | | | | | | |
|---|---|---|---|---|---|---|---|---|
| **Variable** | **Definition** | **Values** | **Field type** | **Source** | **Survey Instruments** | | | |
| | | | | | **EM** | **FU** | **PP 1** | **PP 2** |
| **1. BELpREG record id** | Unique numeric code | Integer (Automatically assigned) | Text box | Automatically assigned | x | x | x | x |
| **2. First name** | Participant’s first name | Text | Text box | *Derived from eConsent* | | | | |
| **3. Last name** | Participant’s last name | Text | Text box | *Derived from eConsent* | | | | |
| **4. E-mail address** | E-mail address used for study participation | Validated text structure (e-mail) | Text box | *Derived from eConsent* | | | | |
| **5. Date of birth** | Date of birth participant | Date (DD-MM-YYYY) | Text box | Reported | x | | | |
| **6. Age** | Age of the participant at first day of the LMP[1] | Integer | Calculated field | *Derived* | | | | |
| **Residence** | | | | | | | | |

| | | | | | |
|---|---|---|---|---|---|
| 7. Country of residence | Country of current residence | Text - selection from drop-down list[2] or free text | Drop-down list linked to database, Autocomplete | Reported | x |
| 8. City of residence | Name and postal code of current residence | Text - selection from drop-down list[3] or free text | Drop-down list linked to database, Autocomplete | Reported | x |
| **9. Partner** | Currently having a partner | Y/N | Radio buttons | Reported | x |
| **10. Father** | Current partner is biological father | Y/N | Radio buttons | Reported | x |
| **11. Year of birth biological father** | Year of birth biological father | YYYY | Text box | Reported | x |
| **Nationality** | | | | | |
| 12. Nationality participant | Nationality participant | Belgian; Dutch; French; German; Moroccan; Romanian; Turkish; Polish; Syrian; Other (specify[2]) | Checkboxes, Multiple choice | Reported | x |
| 13. Nationality biological father | Nationality biological father | Belgian; Dutch; French; German; Moroccan; Romanian; Turkish; Polish; Syrian; Other (specify[2]) | Checkboxes, Multiple choice | Reported | x |
| **Ethnicity** | | | | | |
| 14. Ethnicity participant | Ethnicity of the participant | European; Maghreb; Rest of Africa; Near/Middle East; Pacific/Far East; Rest of Asia; North America; Central and South America; I don't know / I prefer not to say | Checkboxes, Multiple choice | Reported | x |
| 15. Ethnicity biological father | Ethnicity of the biological father | European; Maghreb; Rest of Africa; Near/Middle East; Pacific/Far East; Rest of Asia; North America; Central and South America; I don’t know / I prefer not to say | Checkboxes, Multiple choice | Reported | x |
| **16. Home language** | Language spoken at home | Dutch; French; English; German; Arabic; Turkish; Berber; Other (specify) | Checkboxes, Multiple choice | Reported | x |
| **Education** | | | | | |
| 17. Highest educational level of participant | Highest educational degree obtained by participant | Primary education; Lower secondary education (certificate 2nd degree); Higher secondary education; Higher vocational education (HBO5); Professional Bachelor | Radio buttons | Reported | x |

| | | | | | |
|---|---|---|---|---|---|
| | | / Higher education short type (non-university); Academic Bachelor; Master; PhD; Other (specify) | | | |
| 18. Education in healthcare of participant | A diploma in healthcare obtained by the participant | Y; N; Currently following education in healthcare | Radio buttons | Reported | x |
| 19. Education in healthcare of participant – type | Type of education in healthcare by the participant | Text | Text box | | x |
| 20. Highest educational level of partner | Highest educational degree obtained by current partner | Primary education; Lower secondary education (certificate 2nd degree); Higher secondary education; Higher vocational education (HBO5); Professional Bachelor / Higher education short type (non-university); Academic Bachelor; Master; PhD; Other (specify) | Radio buttons | Reported | x |
| **Profession** | | | | | |
| 21. Professional activity participant | The participant being professional active in the past year | Y/N | Radio buttons | Reported | x |
| 22. Professional activity participant - week hours | Number of hours per week participant was professionally active in the past year | < 20h/week; 20-40h/week; 40-60h/week; >60h/week | Radio buttons | Reported | x |
| 23. Professional activity participant - healthcare | The participant being professional active as healthcare professional in the past year | Y/N | Radio buttons | Reported | x |
| 24. Occupational exposure to substances participant | Occupational exposure to substances by participant in the past year | Y/N | Radio buttons | Reported | x |
| 25. Occupational exposure to substances participant – type | Type of occupational substances to which the participant was exposed to in the past year | Text | Notes box | Reported | x |
| 26. Professional activity partner | The current partner being professional active in the past year | Y/N | Radio buttons | Reported | x |
| **27. Gross Annual Family Income** | Gross Annual Family Income | <15 000 euro; 15 000 euro - <30 000 euro; 30 000 euro - <45 000 euro; 45 000 euro - <65 000 euro; >=65 000 euro; I don't know / prefer not to say | Radio buttons | Reported | x |
| **28. Channel through which the participant was first informed** | Channel through which the participant first heard about the BELpREG study | A healthcare professional informed me (specify type); A friend / family member / neighbour / colleague informed me; Informed by a flyer / poster (specify where); Informed by social media | Checkboxes, Multiple choice | Reported | x |

| | | |
|---|---|---|
| | | (specify type); Informed by media (specify); Informed by an article (specify); Informed by the website apotheek.be; Informed by the website gezondzwangerworden.be; Other (specify) |

Abbreviations : EM = Pregnancy Enrolment Questionnaire; FU = Pregnancy Follow-up Questionnaire; PP 1 = First Postpartum Questionnaire; PP 2 = Second Postpartum Questionnaire
Notes: [1] In case the last menstrual period (LMP) is not directly registered by participants, it is derived from estimated date of delivery (EDD); [2]Structured text field with autocomplete function, linked to NATO database with country names and alpha-2 codes; [3]Structured text field with autocomplete function, linked to the open-source databases of the national post service in Belgium with town names and postal codes

| **Category 2: Current pregnancy and health status** | | | | | | | | |
|---|---|---|---|---|---|---|---|---|
| **Variable** | **Definition** | **Values** | **Field type** | **Source** | **Survey Instruments** | | | |
| | | | | | **EM** | **FU** | **PP 1** | **PP 2** |
| **29. Pregnancy status** | Status of still being pregnant | Y/N | Radio buttons | | | x | | |
| **Gestational age** | | | | | | | | |
| 30. Self-reported gestational age | Self-reported current gestational age | Weeks (integer), days (integer – optional) and 'I don't know (yet)' checkbox | Embedded Text boxes and Checkbox | Reported | x | x | | |
| 31. Calculated gestational age | Gestational age calculated based on EDD[1] | Number | Calculated field | Derived | | | | |
| 32. Gestational age at identification pregnancy | Gestational age at determination pregnancy | Number | Text box | Reported | x | | | |
| **Estimated date of delivery** | | | | | | | | |
| 33. Estimated date of delivery (EDD) | Estimated date of delivery | Date (D-M-Y) and 'I don't know yet' checkbox | Text box and Checkbox | Reported | x | | | |
| 34. Source of the estimated date of delivery | Method used to define the estimated date of delivery | Ultrasound; Calculated based on the first day of the last period; Calculated based on the date of conception (in case of fertility treatment); Other (Specify) | Radio buttons with embedded Text box | Reported | x | | | |
| 35. Last Menstrual Period (LMP) | Date of the first day of the last menstrual period prior to conception | Date (D-M-Y) | Text box | Reported | x | | | |

| | | | | | |
|---|---|---|---|---|---|
| **36. Onset of pregnancy** | Onset of the current pregnancy | Spontaneously; After fertility treatment with hormonal stimulation; After fertility treatment without hormonal stimulation | Radio buttons | Reported | x |
| **Plurality** | | | | | |
| 37. Singleton or multiple pregnancy | Singleton or multiple pregnancy | Singleton; Multiples; I don't know yet | Radio buttons | Reported | x |
| 38. Number of fetuses | Number of fetuses in current pregnancy | Twins; Triplets, Other (specify - integer) | Radio buttons | Reported | x |
| **Pregnancy preparation and planning** | | | | | |
| 39. Planned pregnancy | Planned pregnancy | Y/N | Radio buttons | Reported | x |
| 40. Time until conception | Number of months until conception | Number (+ optional text field to clarify) | Text box | Reported | x |
| 41. Preparation for pregnancy | Preparation for pregnancy | Y/N | Radio buttons | Reported | x |
| 42. Type of preparation for pregnancy | How the participant prepared for the current pregnancy | To avoid infections; To have antibodies checked in blood; To check vaccination status; To pursue ideal weight; To get (physical) exercise; To eat healthier; To discuss the use of medicines with an health care professional; To search for online information; To start folic acid intake | Checkboxes, Multiple choice | Reported | x |
| 43. Expected place of delivery | Expected place of delivery | Hospital; At home; Other (specify); I don't know yet | Radio buttons | Reported | x |
| 44. Expected hospital of delivery | Expected hospital of delivery | Text - selection from drop-down list[2] or free text | Drop-down list linked to database, Autocomplete | Reported | x |
| 45. Willingness to breastfeed | Willingness to breastfeed | Y; N; I don't know | Radio buttons | Reported | x |
| 46. Profession of involved HCPs | Types of HCPs involved in the current pregnancy follow-up | Gynaecologist; Midwife; General Practitioner; Medical specialist (specify type); Psychologist; Other (specify) | Checkboxes, Multiple choice | Reported | x |
| 47. Name(s) of involved HCPs | Name(s) of HCPs involved in the current pregnancy follow-up | Text - selection from drop-down list[3] or free text | Drop-down list linked to database, Autocomplete | Reported | x |
| **Health status participant** | | | | | |

| | | | | | | | |
|---|---|---|---|---|---|---|---|
| 48. Length | Length at conception, in centimeter | Number | Text box | Reported | x | | |
| 49. Weight | Weight at conception, in kilogram | Number | Text box | Reported | x | | |
| 50. Body-Mass index (BMI) | BMI, calculated based on length and weight | Number | Calculated field | Derived | | | |
| 51. Chronic conditions | Chronic conditions (i.e., conditions existing before the start of the pregnancy) | Text - selection from drop-down list[4] or free text - multiple answers possible | Drop-down list linked to database, Autocomplete | Reported | x | | |
| **52. Chronic conditions of the biological father** | Chronic conditions of the biological father, (i.e., existing before the start of the pregnancy) | Text - selection from drop-down list[4] or free text - multiple answers possible - including 'I don't know' option | Drop-down list linked to database, Autocomplete | Reported | x | | |
| **53. Consanguinity** | Mother and biological father have one or more ancestors in common | Y/N | Radio buttons | Reported | x | | |
| **54. Pregnancy complications** | Pregnancy complications (since previous questionnaire) | No complication(s) or disease(s); Severe pregnancy vomiting (hyperemesis gravidarum); Diabetes diagnosed after 20 weeks ('gestational diabetes'); Increased blood pressure after 20 weeks ('Gestational Hypertension'); Pre-eclampsia diagnosed before/during week 34; Pre-eclampsia diagnosed after week 34; HELLP syndrome; Seizures; Growth retardation in the unborn child; Blood clot (thrombosis); Placenta praevia; Placenta abruptio; (severe) bleeding (specify); Depression during pregnancy; Anxiety during pregnancy; Psychosis; Exacerbation of my chronic condition (specify which condition this concerns); Pregnancy cholestasis; Too much amniotic fluid; Too little amniotic fluid; Threatening premature birth; Premature ruptured of membranes (PROM); Premature labour (< 37 weeks); Infection (specify); Other (specify) | Checkboxes with embedded text fields, Multiple choice | Reported | x | x | x |

| | | | | | | | | |
|---|---|---|---|---|---|---|---|---|
| **Maternal hospital admission** | | | | | | | | |
| 55. Hospital admission during pregnancy | Hospital admission since start pregnancy / since previous questionnaire | Y/N | Radio buttons | Reported | x | x | x | |
| 56. Reason for hospital admission during pregnancy | Reason for hospital admission since start pregnancy / since previous questionnaire | Severe pregnancy vomiting (hyperemesis gravidarum); Threatening premature birth; Premature labour (< 37 weeks); Premature rupture of the membranes (PROM); Pre-eclampsia diagnosed before/during week 34; Pre-eclampsia diagnosed after week 34; HELLP syndrome; Seizures; Growth retardation in the unborn child; Blood clot (thrombosis); Placenta praevia; Placental abruptio; (severe) bleeding; Psychosis; Exacerbation of my chronic condition (specify); Infection (specify); Pregnancy cholestasis; Too much amniotic fluid; Too little amniotic fluid; Cardiac arrhythmias; Other (specify) | Checkboxes with embedded text fields, Multiple choice | Reported | x | x | x | |
| **Prenatal screening** | | | | | | | | |
| 57. NIP test | Performance of a non-invasive prenatal test (NIP test) | Y; N; I don't know | Radio buttons | Reported | | | x | |
| 58. Congenital anomaly identified during pregnancy | Identification of a congenital anomaly during pregnancy | Y/N | Radio buttons | Reported | | | x | |
| 59. Source of identification of the congenital anomaly | Source of the identification of the congenital anomaly | NIP test; Ultrasound; Amniocentesis or chorionic villus sampling; Blood test (to determine infections) (specify infection), Other (specify) | Checkboxes, Multiple choice | Reported | | | x | |
| **60. Depressive symptoms** | Edinburgh Depression Scale (EDS-10) | 10 questions with each time 4 response options | Radio buttons (10) | Reported | x | | | x |

Abbreviations : EM = Pregnancy Enrolment Questionnaire; FU = Pregnancy Follow-up Questionnaire; PP 1 = First Postpartum Questionnaire; PP 2 = Second Postpartum Questionnaire; AC = Auto Complete; MC = Multiple Choice; EDD = Estimated Date of Delivery; LMP = Last Menstrual Period; HCPs = Healthcare professionals; BMI = Body-Mass Index; ICD-11 = International Classification of Diseases 11th Revision; MedDRA = Medical Dictionary for Regulatory Activities; HELLP = Haemolysis, Elevated Liver enzymes and Low Platelets.

Notes: [1] If the estimated date of delivery (EDD) is not known, the calculation of gestational age is based on the last menstrual period (LMP); [2] Structured text field with autocomplete function, linked to the open-source database with names and cities of hospitals in Belgium; [3]Structured text field with autocomplete function, linked to the open-source RIZIV/FGOV database with registered HCPs in Belgium, including their name, profession, qualification and work address; [4]Structured text field with autocomplete function, linked to a self-developed list of the most frequently reported disorders linked to their respective ICD-11 and MedDRA classification.

| Category 3: Maternal-obstetric history | | | | | | | | |
|---|---|---|---|---|---|---|---|---|
| **Variable** | **Definition** | **Values** | **Field type** | **Source** | **Survey Instruments** | | | |
| | | | | | **EM** | **FU** | **PP 1** | **PP 2** |
| **61. Previous pregnancy** | Previously having been pregnant | Y/N | Radio buttons | Reported | x | | | |
| **62. Number of previous pregnancies** | Number of previous pregnancies | Integer | Text box | Reported | x | | | |
| **63. Gravidity** | Gravidity | Primigravida; Multigravida | Calculated field | Derived | | | | |
| **History of miscarriage** | | | | | | | | |
| 64. Previous miscarriage | Previous spontaneous interruption of pregnancy before the end of week 22, calculated from the first day of the last menstrual period | Y/N | Radio buttons | Reported | x | | | |
| 65. Number of previous miscarriages | Number previous miscarriages | Integer | Text box | Reported | x | | | |
| 66. Gestational age at previous miscarriage(s) | Gestational age at previous miscarriage(s) | Text | Text box | Reported | x | | | |
| **History of elective termination of pregnancy (ETOP)** | | | | | | | | |
| 67. Previous ETOP | Previous ETOP | Y/N | Radio buttons | Reported | x | | | |
| 68. Number of previous ETOPs | Number of previous ETOPs | Integer | Text box | Reported | x | | | |
| 69. Gestational age at ETOP(s) | Gestational age at previous ETOP(s) | Text | Text box | Reported | x | | | |
| 70. Reason of ETOP(s) | Reason of ETOP(s) | The pregnancy(ies) was (were) terminated due to medical reasons for myself; The pregnancy(ies) was (were) terminated for medical reasons in the embryo/fetus; The pregnancy(ies) was (were) terminated at my personal request | Radio buttons | Reported | x | | | |
| 71. Medical reason of ETOP(s) | The actual medical reason of ETOP(s) | Text | Notes box | Reported | x | | | |
| **History of stillbirth** | | | | | | | | |
| 72. Previous stillbirth | Death of an unborn infant in the womb after 22 weeks of gestation | Y/N | Radio buttons | Reported | x | | | |
| 73. Gestational age at stillbirth(s) | Gestational age at stillbirth(s) | Text | Text box | Reported | x | | | |

| | | | | | |
|---|---|---|---|---|---|
| 74. Congenital anomaly | Identification of a congenital anomaly in the stillborn fetus | Y (specify) / N | Radio buttons with embedded Text box | Reported | x |
| **75. Parity** | Parity | Nullipara, Primipara, Multipara | Calculated field | Derived | |
| **76. History of fertility treatment** | Fertility treatment before the current pregnancy | Y/N | Radio buttons | Reported | x |
| **History of pregnancy / delivery complication(s)** | | | | | |
| 77. History of pregnancy complications | Pregnancy complications during previous pregnancy(ies) | No complication(s); Severe pregnancy vomiting (hyperemesis gravidarum); Diabetes diagnosed after 20 weeks ('gestational diabetes'); Increased blood pressure after 20 weeks ('gestational hypertension'); Pre-eclampsia diagnosed before/during week 34; Pre-eclampsia diagnosed after week 34; HELLP syndrome; Seizures; Growth retardation in the unborn child; Blood clot (thrombosis); Placenta praevia; Placental abruptio; (severe) bleeding (specify); Depression during pregnancy; Anxiety during pregnancy; Psychosis; Exacerbation of my chronic condition (specify which condition this concerns); Pregnancy cholestasis; Too much amniotic fluid; Too little amniotic fluid; Threatening premature birth; Premature rupture of the membranes (PROM); Premature labour (< 37 weeks); Infection (specify); Other (specify) | Checkboxes with embedded text fields, Multiple choice | Reported | x |
| 78. History of delivery complications | Delivery complications during previous pregnancy(ies) | No Complications; Giving birth before 32 weeks; Giving birth between 32 and 37 weeks; New-born with a low birth weight (< 2500 grams); New-born with a high birth weight (>=4000 grams); Blood clot | Checkboxes with embedded text fields, | Reported | x |

| | | | | | |
|---|---|---|---|---|---|
| | | (thrombosis); Coloured amniotic fluid; Shoulder dystocia; No birth of the placenta within 30 minutes after birth of the new-born ('placenta retention'); Ruptured uterus ('uterine rupture'); Oxygen deficiency in the child during childbirth; Amniotic fluid embolism; Sepsis; Acute renal failure; Severe blood loss after delivery (>1000mL); High blood pressure; Cardiac arrhythmias; Postpartum Depression; Anxiety after childbirth; Post-traumatic stress syndrome after childbirth; Psychosis; Other (specify) | Multiple choice | | |
| **Predisposition to congenital anomalies** | | | | | |
| 79. Congenital anomaly during a previous pregnancy | Identification of congenital anomaly(ies) in a previous pregnancy of the participant and/or biological father of the unborn child | Y; N; I don't know | Radio buttons | Reported | x |
| 80. Description of congenital anomaly(ies) in a previous pregnancy | Description of the congenital anomaly(ies) in a previous pregnancy of the participant and/or biological father of the unborn child | Text | Notes box | Reported | x |
| 81. Cause of congenital anomaly(ies) in a previous pregnancy | Cause of the congenital anomaly(ies) in a previous pregnancy of the participant and/or biological father of the unborn child | There is no known genetic cause; There is a known genetic cause; A suspected genetic cause is known; I don't know; Other (specify) | Checkboxes, Multiple choice | Reported | x |
| 82. Congenital anomaly in the participant | Identification of congenital anomaly(ies) in the participant | Y/N | Radio buttons | Reported | x |
| 83. Description of congenital anomaly(ies) in the participant | Description of the congenital anomaly(ies) in the participant | Text | Notes box | Reported | x |
| 84. Cause of congenital anomaly(ies) in the participant | Cause of the congenital anomaly(ies) in the participant | There is no known genetic cause; There is a known genetic cause; A suspected genetic cause is known; Other (specify) | Checkboxes, Multiple choice | Reported | x |
| 85. Congenital anomaly in the father | Identification of congenital anomaly(ies) in the biological father | Y; N; I don't know | Radio buttons | Reported | x |
| 86. Description of congenital anomaly(ies) in the father | Description of the congenital anomaly(ies) in the biological father | Text | Notes box | Reported | x |

| | | | | | |
|---|---|---|---|---|---|
| 87. Cause of congenital anomaly(ies) in the father | Cause of the congenital anomaly(ies) in the biological father | There is no known genetic cause; There is a known genetic cause; A suspected genetic cause is known; I don't know; Other (specify) | Checkboxes, Multiple choice | Reported | x |
| 88. Congenital anomaly(ies) in a parent or sibling | Identification of congenital anomaly(ies) in parent or sibling of the participant and/or biological father | Y; N; I don't know | Radio buttons | Reported | x |
| 89. Description of congenital anomaly(ies) in a parent or sibling | Description of congenital anomaly(ies) in a parent or sibling of mother or biological father, including in whom this was diagnosed | Text | Notes box | Reported | x |
| 90. Cause of congenital anomaly(ies) in a parent or sibling | Cause of congenital anomaly(ies) in a parent or sibling of mother or biological father | There is no known genetic cause; There is a known genetic cause; A suspected genetic cause is known; I don't know; Other (specify) | Checkboxes, Multiple choice | Reported | x |
| **Predisposition to developmental disorders** | | | | | |
| 91. Developmental disorder during previous pregnancy | Identification of developmental disorder(s) in a previous pregnancy of the participant and/or the biological father of the unborn child | Y; N; I don't know | Radio buttons | Reported | x |
| 92. Description developmental disorder(s) during previous pregnancy | Description of developmental disorder(s) in a previous pregnancy of the participant and/or the biological father of the unborn child | Text | Notes box | Reported | x |
| 93. Developmental disorder in the participant | Identification of developmental disorder(s) in the participant | Y/N | Radio buttons | Reported | x |
| 94. Description of developmental disorder(s) in the participant | Description of developmental disorder(s) in the participant | Text | Notes box | Reported | x |
| 95. Developmental disorder in the father | Identification of developmental disorder(s) in the biological father | Y; N; I don't know | Radio buttons | Reported | x |
| 96. Description of developmental disorder(s) in the father | Descriptions of developmental disorder(s) in the biological father | Text | Notes box | Reported | x |

| | | | | | |
|---|---|---|---|---|---|
| 97. Developmental disorder in a parent or sibling | Identification of developmental disorder(s) in a parent or sibling of mother or biological father | Y; N; I don't know | Radio buttons | Reported | x |
| 98. Description of developmental disorder(s) in a parent or sibling | Descriptions of developmental disorder(s) in a parent or sibling of mother or biological father, including the notification in whom this was diagnosed | Text | Notes box | Reported | x |
| **Predisposition to motor disorders** | | | | | |
| 99. Motor disorder during a previous pregnancy | Identification of motor disorder(s) in a previous pregnancy of the participant and/or biological father of the unborn child | Y; N; I don't know | Radio buttons | Reported | x |
| 100. Description motor disorder(s) during a previous pregnancy | Description of motor disorder(s) in a previous pregnancy of the participant and/or the biological father of the unborn child | Text | Notes box | Reported | x |
| 101. Motor disorder in the participant | Identification of motor disorder(s) in the participant | Y/N | Radio buttons | Reported | x |
| 102. Description motor disorder(s) in the participant | Description of motor disorder(s) in the participant | Text | Notes box | Reported | x |
| 103. Motor disorder in the father | Identification of motor disorder(s) in the biological father | Y; N; I don't know | Radio buttons | Reported | x |
| 104. Description motor disorder(s) in the father | Description of motor disorder(s) in the biological father | Text | Notes box | Reported | x |

Abbreviations : EM = Pregnancy Enrolment Questionnaire; FU = Pregnancy Follow-up Questionnaire; PP 1 = First Postpartum Questionnaire; PP 2 = Second Postpartum Questionnaire; ETOP = Elective Termination Of Pregnancy; HELLP = Haemolysis, Elevated Liver enzymes and Low Platelets.

| **Category 4: Use of medicines, folic acid / pregnancy vitamins and other health products** | | | | | | | | |
|---|---|---|---|---|---|---|---|---|
| **Variable** | **Definition** | **Values** | **Field type** | **Source** | **Survey Instruments** | | | |
| | | | | | **EM** | **FU** | **PP 1** | **PP 2** |
| **Medication use during pregnancy** - The following questions are repeated for each reported medicine | | | | | | | | |
| **105. Name of the medicine** | Name of the medicine used since the start of the pregnancy (i.e., in the enrolment questionnaire) or since completing the previous questionnaire (i.e., in a follow-up questionnaire) or since completing the previous questionnaire and the end of the pregnancy (i.e., in the postpartum 1 questionnaire) | Text - selection from drop-down list[1] or free text | Drop-down list linked to database, Autocomplete | Reported | x | x | x | |
| **106. Modality of medicine use** | How was/has the medicine been used | a) Daily and still (e.g., thyroid hormone);<br>b) Occasionally or as needed (e.g., a painkiller taken occasionally, or an injection that is regularly given);<br>c) During a certain period and not anymore (e.g., an antibiotic that was used for 5 days) | Radio buttons | Reported | x | x | x | |
| ***Medication use during pregnancy – modality 'a) Daily and still'*** | | | | | | | | |
| 107. How many times a day | How many times the medicine has been used a day | 1x per day; 2x a day; 3x per day; Other (specify) | Radio buttons | Reported | x | x | x | |
| 108. Quantity per intake – number | Number of units per intake | ½; 1; 2; 3; 4; Other (specify) | Text box | Reported | x | x | x | |
| 109. Quantity per intake – unit | Unit of intake | Capsule(s) or tablet(s); Teaspoon(s) (5mL); Tablespoon(s) (15mL); Spray(s) (e.g., per nostril); Puff(s); Drop(s) (e.g., per nostril or eye); Syringe(s) / ampoule(s); Units; Suppository(s) / ovule(s); Fingertip(s) (with ointment/cream/gel); Patch(s); Sachet(s); Other (specify) | Dropdown | Reported | x | x | x | |
| 110. Start of use | Timing of medication use initiation | Before this pregnancy; During this pregnancy | Radio buttons | Reported | x | | | |

| | | | | | | | |
|---|---|---|---|---|---|---|---|
| 111. Start of use during pregnancy – date | Start date of medication use during this pregnancy | Date (DD-MM-YYYY) | Text box | Reported | x | x | x |
| 112. Continuation of use | Continuation of a previously registered medicine under the modality 'Daily and still'[3]. | Yes; I no longer use this medicine (if so, when did you stop); The use of this medicine has changed (if so, date of change and variables 107-109 are questioned) | Radio buttons | Reported | | x | x |
| ***Medication use during pregnancy - modality 'b) Occasionally or as needed'*** | | | | | | | |
| 113. Number of days when medicine was used | Number of days when the medicine was used since the start of the pregnancy (i.e., in the enrolment questionnaire) or since completing the previous questionnaire (i.e., in a follow-up questionnaire) or since completing the previous questionnaire and the end of the pregnancy (i.e., in the postpartum 1 questionnaire) | - During the first trimester (from week 1 to the end of week 12): integer<br>- During the second trimester (from week 13 to the end of week 26): integer<br>- During the third trimester (from week 27 until the end of the pregnancy): integer | Text box (3) | Reported | x | x | x |
| 114. How many times a day | How many times the medicine was used a day | 1x per day; 2x a day; 3x per day; Variable (specify); Other (specify) | Radio buttons | Reported | x | x | x |
| 115. Quantity per intake - number | Number of units per intake | 1/2; 1; 2; 3; 4; Other (specify) | Text box | Reported | x | x | x |
| 116. Quantity per intake - unit | Unit of intake | Capsule(s) or tablet(s); Teaspoon(s) (5mL); Tablespoon(s) (15mL); Spray(s) (e.g., per nostril); Puff(s); Drop(s) (e.g., per nostril or eye); Syringe(s) / ampoule(s); Units; Suppository(s) / ovule(s); Fingertip(s) (with ointment/cream/gel); Patch(s); Sachet(s); Other (specify) | Dropdown | Reported | x | x | x |
| 117. Start of use during pregnancy - date | Start date of medication use during this pregnancy | Date (DD-MM-YYYY) | Text box | Reported | x | x | x |
| 118. Use in 6 months before start pregnancy | Use of the medicine in the 6 months before the start of the pregnancy | Y/N | Radio buttons | Reported | x | | |
| ***Medication use during pregnancy - modality 'c) During a certain period and not anymore'*** | | | | | | | |
| 119. How many times a day | How many times the medicine was used a day | 1x per day; 2x a day; 3x per day; Variable (specify); Other (specify) | Radio buttons | Reported | x | x | x |
| 120. Quantity per intake - number | Number of units per intake | 1/2; 1; 2; 3; 4; Other (specify) | Text box | Reported | x | x | x |

| | | | | | | | |
|---|---|---|---|---|---|---|---|
| 121. Quantity per intake - unit | Unit of intake | Capsule(s) or tablet(s); Teaspoon(s) (5mL); Tablespoon(s) (15mL); Spray(s) (e.g., per nostril); Puff(s); Drop(s) (e.g., per nostril or eye); Syringe(s) / ampoule(s); Units; Suppository(s) / ovule(s); Fingertip(s) (with ointment/cream/gel); Patch(s); Sachet(s); Other (specify) | Dropdown | Reported | x | x | x |
| 122. Start of use | Timing of medication use initiation | Before this pregnancy; During this pregnancy | Radio buttons | Reported | x | x | x |
| 123. Start of use during pregnancy - date | Start date of medication use during this pregnancy | Date (DD-MM-YYYY) | Text box | Reported | x | x | x |
| 124. Stop of use | Stop date of medication use | Date (DD-MM-YYYY) | Text box | Reported | x | x | x |
| **125. Indication of medication use** | Indication of the medication use, as reported by the participant | Text - selection from drop-down list[3] or free text | Drop-down list linked to database, Autocomplete | Reported | x | x | x |
| **4.2 Use of folic acid and pregnancy vitamins during pregnancy** - The following questions are repeated for each reported folic acid product and pregnancy vitamin | | | | | | | |
| **126. Name of the folic acid product or pregnancy vitamin** | Name of the folic acid product or pregnancy vitamin used currently (i.e., in the enrolment questionnaire) or since completing the previous questionnaire (i.e., in a follow-up questionnaire) or since completing the previous questionnaire and the end of the pregnancy (i.e., in the postpartum 1 questionnaire) | Text – selection from drop-down list[1] or free text | Drop-down list linked to database, Autocomplete | Reported | x | x | x |
| 127. Start of use | Start date of using the folic acid product or pregnancy vitamin | Date (DD-MM-YYYY) | Text box | Reported | x | x | x |
| 128. How many times a day | How many times the folic acid product or pregnancy vitamin was/has been used | 1x per day; Other (specify) | Radio buttons | Reported | x | x | x |
| 129. Stop of use | Stop date of using the folic acid product or pregnancy vitamin | Date (DD-MM-YYYY) | Text box | Reported | | x | x |
| 130. Continuation of use | Continuation of a previously registered folic acid product or pregnancy vitamin[4] | Yes; I no longer use this product (if so, when did you stop); The use of this | Radio buttons | Reported | | x | x |

| | | | | | | | |
|---|---|---|---|---|---|---|---|
| | | product has changed (if so, date of change and variable 128 is questioned) | | | | | |
| **131. Name of a folic acid product or pregnancy vitamin used earlier in pregnancy** | Name of a folic acid product or pregnancy vitamin used earlier in this pregnancy | Text - selection from drop-down list[1] or free text | Drop-down list linked to database, Autocomplete | Reported | x | | |
| 132. How many times a day | How many times the folic acid product or pregnancy vitamin was used | 1x per day; Other (specify) | Radio buttons | Reported | x | | |
| 133. Start of use | Start date of using the folic acid product or pregnancy vitamin | Date (DD-MM-YYYY) | Text box | Reported | x | | |
| 134. Stop of use | Stop date of using the folic acid product or pregnancy vitamin | Date (DD-MM-YYYY) | Text box | Reported | x | | |
| **4.3 Health products' use during pregnancy** - The following questions are repeated for each reported health product | | | | | | | |
| **135. Name of the health product** | Name of the health product used since the start of the pregnancy (i.e., in the enrolment questionnaire) or since completing the previous questionnaire (i.e., in a follow-up questionnaire) or since completing the previous questionnaire and the end of the pregnancy (i.e., in postpartum 1 questionnaire) | Text - selection from drop-down list[1] or free text | Drop-down list linked to database, Autocomplete | Reported | x | x | x |
| **136. Modality of health product use** | How was/has the health product been used | a) Daily and still (e.g., thyroid hormone);<br>b) Occasionally or as needed (e.g., a painkiller taken occasionally, or an injection that is given regularly);<br>c) During a certain period and not anymore (e.g., an antibiotic that was used for 5 days) | Radio buttons | Reported | x | x | x |
| ***Health products' use during pregnancy - modality 'a) Daily and still'*** | | | | | | | |
| 137. How many times a day | How many times the health product has been used a day | 1x per day; 2x a day; 3x per day; Other (specify) | Radio buttons | Reported | x | x | x |
| 138. Quantity per intake - number | Number of units per intake | 1/2; 1; 2; 3; 4; Other (specify) | Text box | Reported | x | x | x |
| 139. Quantity per intake - unit | Unit of intake | Capsule(s) or tablet(s); Teaspoon(s) (5mL); Tablespoon(s) (15mL); Spray(s) (e.g., per nostril); Puff(s); Drop(s) (e.g., per nostril or eye); Syringe(s) / ampoule(s); Units; Suppository(s) / | Dropdown | Reported | x | x | x |

| | | | | | | | |
|---|---|---|---|---|---|---|---|
| | | ovule(s); Fingertip(s) (with ointment/cream/gel); Patch(s); Sachet(s); Other (specify) | | | | | |
| 140. Start of use | Timing of the initiation of the use of the health product | Before this pregnancy; During this pregnancy | Radio buttons | Reported | x | | |
| 141. Start of use during pregnancy - date | Start date of using the health product during this pregnancy | Date (DD-MM-YYYY) | Text box | Reported | x | x | x |
| 142. Continuation of use | Continuation of a previously registered health product under the modality 'Daily and still'[2]. | Yes; I no longer use this product (if so, when did you stop); The use of this product has changed (if so, date of change and variables 137-139 is questioned) | Radio buttons | Reported | | x | x |
| ***Health products' use during pregnancy - modality 'b) Occasionally or as needed'*** | | | | | | | |
| 143. Number of days when heath product used | Number of days when the health product was used since the start of the pregnancy (i.e., in the enrolment questionnaire) or since completing the previous questionnaire (i.e., in a follow-up questionnaire) or since completing the previous questionnaire and the end of the pregnancy (i.e., in the postpartum 1 questionnaire) | - During the first trimester (from week 1 to the end of week 12): integer<br>- During the second trimester (from week 13 to the end of week 26): integer<br>- During the third trimester (from week 27 until the end of the pregnancy): integer | Text box (3) | Reported | x | x | x |
| 144. How many times a day | How many times the health product was used a day | 1x per day; 2x a day; 3x per day; Variable (specify); Other (specify) | Radio buttons | Reported | x | x | x |
| 145. Quantity per intake - number | Number of units per intake | 1/2; 1; 2; 3; 4; Other (specify) | Text box | Reported | x | x | x |
| 146. Quantity per intake - unit | Unit of intake | Capsule(s) or tablet(s); Teaspoon(s) (5mL); Tablespoon(s) (15mL); Spray(s) (e.g., per nostril); Puff(s); Drop(s) (e.g., per nostril or eye); Syringe(s) / ampoule(s); Units; Suppository(s) / ovule(s); Fingertip(s) (with ointment/cream/gel); Patch(s); Sachet(s); Other (specify) | Dropdown | Reported | x | x | x |
| 147. Start of use during pregnancy - date | Start date of the use of the health product during this pregnancy | Date (DD-MM-YYYY) | Text box | Reported | x | x | x |

| | | | | | | | |
|---|---|---|---|---|---|---|---|
| 148. Use in 6 months before start pregnancy | Use of the health product in the 6 months before the start of the pregnancy | Y/N | Radio buttons | Reported | x | | |
| ***Health products' use during pregnancy - modality 'c) During a certain period and not anymore'*** | | | | | | | |
| 149. How many times a day | How many times the health product was used a day | 1x per day; 2x a day; 3x per day; Variable (specify); Other (specify) | Radio buttons | Reported | x | x | x |
| 150. Quantity per intake - number | Number of units per intake | 1/2; 1; 2; 3; 4; Other (specify) | Text box | Reported | x | x | x |
| 151. Quantity per intake - unit | Unit of intake | Capsule(s) or tablet(s); Teaspoon(s) (5mL); Tablespoon(s) (15mL); Spray(s) (e.g., per nostril); Puff(s); Drop(s) (e.g., per nostril or eye); Syringe(s) / ampoule(s); Units; Suppository(s) / ovule(s); Fingertip(s) (with ointment/cream/gel); Patch(s); Sachet(s); Other (specify) | Dropdown | Reported | x | x | x |
| 152. Start of use | Timing of the initiation of the use of the health product | Before this pregnancy; During this pregnancy | Radio buttons | Reported | x | x | x |
| 153. Start of use during pregnancy - date | Start date of the use of the health product during this pregnancy | Date (DD-MM-YYYY) | Text box | Reported | x | x | x |
| 154. Stop of use | Stop date of the use of the health product | Date (DD-MM-YYYY) | Text box | Reported | x | x | x |
| **155. Indication of the health product** | Indication of the use of the health product, as reported by the participant | Text - selection from drop-down list[3] or free text | Drop-down list linked to database, Autocomplete | Reported | x | x | x |
| **Other medicines and health products used in the 6 months before the start of the pregnancy** | | | | | | | |
| **156. Name of the medicine or health product** | Name of the medicine or health product used in the 6 months before the start of the pregnancy | Text - selection from drop-down list[1] or free text | Drop-down list linked to database, Autocomplete | Reported | x | | |
| 157. Stop of use | Stop date of the use of the medicine or health product (estimation) | Date (DD-MM-YYYY) | Text box | Reported | x | | |
| **Paternal medication use** | | | | | | | |
| **158. Name of the medicine or health product** | Name of the medicine or health product used by the biological father in the 3 months before the start of the pregnancy | Text - selection from drop-down list[1] or free text - including 'I don't know' option | Drop-down list linked to database, Autocomplete | Reported | x | | |
| **Medication use postpartum** | | | | | | | |

| | | | | | | | | |
|---|---|---|---|---|---|---|---|---|
| **159. Name of the medicine or health product** | Name of the medicine or health product used since childbirth | Text - selection from drop-down list[1] or free text | Drop-down list linked to database, Autocomplete | Reported | | | x | x |

Abbreviations : EM = Pregnancy Enrolment Questionnaire; FU = Pregnancy Follow-up Questionnaire; PP 1 = First Postpartum Questionnaire; PP 2 = Second Postpartum Questionnaire; ATC = Anatomical Therapeutic Chemical; CNK = National Code Number; VMP = Virtual Medicinal Product; AMP = Actual Medicinal Product; NMP = Non Medicinal Product; ICD-11 = International Classification of Diseases 11th Revision; MedDRA = Medical Dictionary for Regulatory Activities.

Notes: [1]Structured text field with autocomplete function, including pictures of the products, linked to a database of available medicines and medicinal products in Belgium. Additional information of the product is derived from the linked database, including ATC code, CNK, VMP, VMP group, Substance, Substance strength, Route of drug administration, Pharmaceutical form, Medication / non-medication status; [2]If a medicine or health product was registered as 'Daily and still' in the previous questionnaire, this medicine will be shown with the reported details of use in the next questionnaire. This question is always shown first, after which other products can be registered; [3]Structured text field with autocomplete function, linked to a self-developed list of the most frequent reported indications / conditions that are linked to their respective ICD-11 and MedDRA classification; [4]If a folic acid product or pregnancy vitamin was registered in the previous questionnaire, this medicine will be shown with the reported details of use in the next questionnaire. This question is always shown first, after which other products can be registered.

| **Category 5: Substance use** | | | | | | | | |
|---|---|---|---|---|---|---|---|---|
| **Variable** | **Definition** | **Values** | **Field type** | **Source** | **Survey Instruments** | | | |
| | | | | | **EM** | **FU** | **PP 1** | **PP 2** |
| **Smoking** | | | | | | | | |
| 160. Smoking - ever | Smoking, ever done (normal cigarette or e-cigarette) | Y/N | Radio buttons | Reported | x | | | |
| 161. Smoking - past year | Smoking in the past year | Y/N | Radio buttons | Reported | x | | | |
| 162. Smoking - in pregnancy | Smoking during this pregnancy | Y/N | Radio buttons | Reported | x | | | |
| 163. Timing of smoking cessation | Timing of smoking cessation | In the past year; Since trying to get pregnant; Since I knew I was pregnant | Radio buttons | Reported | x | | | |
| 164. Smoking status throughout pregnancy | Smoking status throughout pregnancy | Y/N | Radio buttons | Reported | x | x | | |
| 165. Timing of smoking cessation in pregnancy | Gestational age upon smoking cessation | Number | Text box | Reported | x | x | | |
| 166. Amount of smoking in pregnancy | Daily amount of smoking in this pregnancy | 1-10 cigarettes/day; 11-20 cigarettes/day; 21-30 cigarettes/day; >30 cigarettes/day | Radio buttons | Reported | x | x | | |
| 167. Postpartum smoking status | Smoking since delivery | Y/N | Radio buttons | Reported | | | x | x |

| | | | | | | | | |
|---|---|---|---|---|---|---|---|---|
| 168. Frequency of smoking since delivery | Frequency of smoking since delivery | 1-10 cigarettes/day; 11-20 cigarettes/day; 21-30 cigarettes/day; >30 cigarettes/day | Radio buttons | Reported | | | x | x |
| 169. Exposure to second-hand smoking during pregnancy | Smoking by someone who lives together with the pregnant participant | Y/N | Radio buttons | Reported | x | | | |
| 170. Amount of second-hand smoking during pregnancy | Daily amount of smoking by someone who lives together with the pregnant participant | 1-10 cigarettes/day; 11-20 cigarettes/day; 21-30 cigarettes/day; >30 cigarettes/day | Radio buttons | Reported | x | | | |
| 171. Second-hand smoking in-house during pregnancy | Smoking in-house by someone who lives together with the pregnant participant | Y/N | Radio buttons | Reported | x | | | |
| **Alcohol use** | | | | | | | | |
| 172. Alcohol use - past year | Use of alcohol in the past year | Y/N | Radio buttons | Reported | x | | | |
| 173. Alcohol use - in pregnancy | Use of alcohol during this pregnancy | Y/N | Radio buttons | Reported | x | | | |
| 174. Timing of alcohol cessation | Timing of alcohol cessation | In the past year; Since trying to get pregnant; Since I knew I was pregnant | Radio buttons | Reported | x | | | |
| 175. Alcohol use status throughout pregnancy | Alcohol use status throughout pregnancy | Y/N | Radio buttons | Reported | x | x | | |
| 176. Timing of alcohol use cessation in pregnancy | Gestational age upon alcohol use cessation | Number | Text box | Reported | x | x | | |
| 177. Alcohol use after delivery | Alcohol use after delivery | Y/N | Radio buttons | Reported | | | x | x |
| 178. Frequency of drinking alcohol in pregnancy | Frequency of drinking alcohol in this pregnancy | Monthly or less; 2-4 times a month; 2-3 times a week; 4 or more times a week | Radio buttons | Reported | x | x | | |
| 179. Frequency of drinking alcohol after delivery | Frequency of drinking alcohol after delivery | Monthly or less; 2-4 times a month; 2-3 times a week; 4 or more times a week | Radio buttons | Reported | | | x | x |
| 180. Amount of alcohol per occasion | Number of standard glasses of alcohol used per occasion. 1 standard glass is the amount of a drink that is usually served in a pub or at a restaurant, e.g., 25cl beer or 10cl wine | <1; 1 or 2; 3 or 4; 5 or 6; 7 or 9; 10 or more | Radio buttons | Reported | x | x | x | x |
| 181. Occurrence of binge drinking | Occurrence of using 6 or more standard glasses of alcohol per occasion | Never; Less than monthly; Monthly; Weekly; Daily or almost daily | Radio buttons | Reported | x | x | x | x |
| 182. Alcohol use in the close environment during pregnancy | Daily use of alcohol by someone who lives together with the pregnant participant | Y/N | Radio buttons | Reported | x | | | |
| **Cannabis and other illicit drugs** | | | | | | | | |

| | | | | | | | | |
|---|---|---|---|---|---|---|---|---|
| 183. Cannabis and other illicit drug use - ever | Cannabis and other illicit drug use, ever used | Y/N | Radio buttons | Reported | x | | | |
| 184. Cannabis and other illicit drug use - past year | Use of cannabis and other illicit drugs in the past year | Y/N | Radio buttons | Reported | x | | | |
| 185. Cannabis and other illicit drug use - in pregnancy | Use of cannabis and other illicit drugs in this pregnancy | Y/N | Radio buttons | Reported | x | | | |
| 186. Timing of cessation of use of cannabis and other illicit drugs | Timing of cessation of the use of cannabis and other illicit drugs | In the past year; Since trying to get pregnant; Since I knew I was pregnant | Radio buttons | Reported | x | | | |
| 187. Type of drugs used | Type of drugs used in pregnancy or after delivery, i.e., cannabis (hasj, weed, marihuana), XTC (MDMA), amphetamines (speed,…), hallucinogens (LSD, magic mushrooms), cocaine, ketamine, heroine, GHB, other (specify) | Never used; Less than once a month; Multiple times a month; Once a week; Multiple times a week; Daily | Matrix with radio buttons | Reported | x | x | x | x |
| 188. Drug use in the close environment during pregnancy | Drug use by someone who lives together with the pregnant participant | Y/N | Radio buttons | Reported | x | | | |

Abbreviations : EM = Pregnancy Enrolment Questionnaire ; FU = Pregnancy Follow-up Questionnaire ; PP 1 = First Postpartum Questionnaire ; PP 2 = Second Postpartum Questionnaire.

| **Category 6: Pregnancy outcomes** | | | | | | | | |
|---|---|---|---|---|---|---|---|---|
| **Variable** | **Definition** | **Values** | **Field type** | **Source** | **Survey Instruments** | | | |
| | | | | | **EM** | **FU** | **PP 1** | **PP 2** |
| **189. Pregnancy outcome** | Identification of the outcome of the current pregnancy | - I gave birth to a live birth child/children;<br>- I had a miscarriage (i.e., a spontaneous termination of pregnancy before the end of pregnancy week 22);<br>- I had an ectopic pregnancy;<br>- My pregnancy was terminated for medical reasons or at my request;<br>- The fetus has died in the womb, after 22 weeks gestational age (i.e., stillbirth) | Checkboxes, Multiple choice | Reported | | | x | |
| ***In case of 'live birth child(ren)'*** | | | | | | | | |

| | | | | | | | |
|---|---|---|---|---|---|---|---|
| **Plurality**[1] | | | | | | | |
| 190. Singleton or multiple pregnancy | Singleton or multiple pregnancy | Singleton; Multiple | Radio buttons | Reported | | | x |
| 191. Number of children | Number of children born after the current pregnancy | Twins; Triplets, Other (specify - integer) | Radio buttons | Reported | | | x |
| **Delivery** | | | | | | | |
| 192. Actual date of delivery | Date of delivery | Date (D-M-Y) | Text box | Reported | | | x |
| 193. Gestational age at delivery | Gestational age at delivery | Weeks (integer) and days (integer) | Text box | Reported | | | x |
| 194. Expected date of delivery | Expected date of delivery | Date (D-M-Y) | Text box | Reported | | | x |
| 195. Mode of delivery | Mode of delivery | Spontaneous vaginal delivery; Assisted (instrumental) vaginal delivery; Planned caesarean section; Unplanned caesarean section; Other (specify) | Radio buttons | Reported | | | x |
| 196. Onset of labor | Onset / start of labor | Spontaneous; Initiated (+reason); Not (yet) in labor at the time of the planned caesarean section; Other (specify) | Radio buttons | Reported | | | x |
| 197. Reason of caesarean section | Reason of the caesarean section | Previous caesarean section; Non-progressive delivery; Congenital anomaly of the neonate (e.g., heart problem, spina bifida...); Fetal distress (e.g., poor heart tones) (specify problems); Medical condition of the pregnant woman (e.g., preeclampsia) (specify problems); Extreme prematurity; Abnormal position of the baby; Placenta praevia; Other (specify) | Checkboxes with embedded text fields, Multiple choice | Reported | | | x |
| 198. Complications after delivery | Complications or medical problems that occurred in the first 72 hours after delivery | No complications; Meconium stained amniotic fluid; Shoulder dystocia; Severe postpartum blood loss (>1000mL); Retained placenta; Uterine rupture; Oxygen deficiency in the neonate during childbirth; Amniotic fluid embolism; Infection (specify type); Sepsis; Acute renal failure; Thrombosis; High blood pressure; Cardiac | Checkboxes with embedded text fields, Multiple choice - Notes box | Reported | | | x |

| | | | | | | | | | |
|---|---|---|---|---|---|---|---|---|---|
| | | arrhythmias; Psychosis; Other (specify) - Including possibility to add free text | | | | | | | |
| 199. Maternal blood transfusion | Maternal blood transfusion after delivery | Y/N | Radio buttons | Reported | | | | x | |
| 200. Admission to a maternal intensive care unit | Admission of the mother to an intensive care unit after delivery | Y/N | Radio buttons | Reported | | | | x | |
| 201. Reason of admission to a maternal intensive care unit | Reason for the admission of the mother to a maternal intensive care unit after delivery | Text | Notes box | Reported | | | | x | |
| 202. Place of delivery | Place of delivery | Hospital; At home; Other (specify) | Radio buttons | Reported | | | | x | |
| 203. Hospital of delivery | City of the hospital of delivery | Text - selection from drop-down list[2] or free text | Drop-down list linked to database, Autocomplete | Reported | | | | x | |
| **Postpartum complications** | | | | | | | | | |
| 204. Postpartum complications | Maternal complications/medical problems that occurred in the first 6 weeks postpartum | No complications; (severe) bleeding (specify); Retained placenta; High blood pressure; Thrombosis/embolism; Mastitis; Breast abscess; Endometritis; Urinary tract infection; Wound problems/infection; Sepsis; Postnatal depression; Psychosis; Flare up of my chronic condition (specify which condition this concerns); Other (specify) - Including possibility to add free text | Checkboxes with embedded text fields, Multiple choice - Notes box | Reported | | | | x | x |
| 205. Maternal hospital admission postpartum | Hospital admission of the mother in the first 6 weeks postpartum | Y/N | Radio buttons | Reported | | | | x | x |
| 206. Reason of maternal hospital admission postpartum | Reason of hospital admission of the mother in the first 6 weeks postpartum | No complications; (severe) bleeding (specify); Retained placenta; High blood pressure; Thrombosis/embolism; Mastitis; Breast abscess; Endometritis; Urinary tract infection; Wound problems/infection; Sepsis; Postnatal depression; Psychosis; Flare up of my chronic condition (specify which | Checkboxes with embedded text fields, Multiple choice - Notes box | Reported | | | | x | x |

| | | | | | | | |
|---|---|---|---|---|---|---|---|
| | | condition this concerns); Other (specify) - Including possibility to add free text | | | | | |
| ***In case of a 'miscarriage'*** | | | | | | | |
| **207. Gestational age at miscarriage** | Gestational age at miscarriage | Weeks (text) | Text box | Reported | | | x |
| **208. Plurality** | Singleton or multiple pregnancy | Y/N | Radio buttons | Reported | | | x |
| **209. Reason of the miscarriage** | Indication of a possible reason of the miscarriage | Y/N | Radio buttons | Reported | | | x |
| **210. Description of the reason of the miscarriage** | Description of the possible reason of the miscarriage | Text | Notes box | Reported | | | x |

| | | | | | | | |
|---|---|---|---|---|---|---|---|
| ***In case of an 'elective termination of pregnancy (ETOP)'*** | | | | | | | |
| **211. Gestational age at ETOP** | Gestational age at ETOP | Weeks (text) | Text box | Reported | | | x |
| **212. Reason of the ETOP** | Reason of the ETOP | The pregnancy was terminated due to maternal medical reasons; The pregnancy was terminated due to medical reasons in the embryo/fetus; The pregnancy was terminated at my personal request | Radio buttons | Reported | | | x |
| **213. Medical reason ETOP** | Medical reason of the ETOP | Text | Notes box | Reported | | | x |
| ***In case of a 'stillbirth'*** | | | | | | | |
| **214. Gestational age at stillbirth** | Gestational age at stillbirth | Text | Text box | Reported | | | x |
| **215. Identification of a congenital anomaly** | Presence of a congenital anomaly in the stillborn fetus | Y (specify) / N | Radio buttons with embedded Text box | Reported | | | x |
| **216. Name of the involved doctor(s) or midwife(s)** ***(asked to all participants)*** | Name of the doctor(s) and/or midwife(s) involved in the pregnancy follow-up | Text - selection from drop-down list[3] or free text | Drop-down list linked to database, Autocomplete | Reported | | | x |

Abbreviations : EM = Pregnancy Enrolment Questionnaire; FU = Pregnancy Follow-up Questionnaire; PP 1 = First Postpartum Questionnaire; PP 2 = Second Postpartum Questionnaire; ETOP = Elective termination Of Pregnancy; NIP = Non-Invasive Prenatal Screening

Notes: [1]In case of a multiple pregnancy, the neonatal outcome fields are multiplied (see category 7); [2]Structured text field with autocomplete function, linked to the open-source hospital database with names and cities of hospitals in Belgium; [3]Structured text field with autocomplete function, linked to the open-source RIZIV/FGOV database with registered HCPs in Belgium, including their name, profession, qualification and work address.

| Category 7: Neonatal outcomes[1] | | | | | | | | |
|---|---|---|---|---|---|---|---|---|
| **Variable** | **Definition** | **Values** | **Field type** | **Source** | **Survey Instruments** | | | |
| | | | | | **EM** | **FU** | **PP 1** | **PP 2** |
| **Biometry at birth** | | | | | | | | |
| 217. Birth weight | Birth weight in gram | Number | Text box | Reported | | | x | |
| 218. Birth length | Birth length in centimeter | Number | Text box | Reported | | | x | |
| 219. Head circumference | Head circumference of the infant at birth | Number - including 'I don't know option' | Text box and Checkbox | Reported | | | x | |
| **220. Sex** | Infant sex | Male; Female; Other (specify) | Text box | Reported | | | x | |
| **221. Apgar score** | Apgar score after 1, 5 and 10 minutes | Integer | Text box (3) | Reported | | | x | |
| **222. Nationality infant** | Nationality infant | Belgian; Dutch; French; German; Moroccan; Romanian; Turkish; Polish; Syrian; Other (specify[2]) | Checkboxes, Multiple choice | Reported | | | x | |
| **Congenital anomalies** | | | | | | | | |
| 223. Congenital anomaly | Identification of a congenital anomaly | Y/N | Radio buttons | Reported | | | x | x |
| 224. Congenital anomaly - Description | Description of the congenital anomaly | Text | Text box | Reported | | | x | x |
| 225. Congenital anomaly - cause | Possible cause of the congenital anomaly | There is no known genetic cause; There is a known genetic cause; A suspected genetic cause is known; I don't know | Checkboxes | Reported | | | x | x |
| 226. Congenital anomaly - surgery | Surgery already occurred or needed in the future due to the congenital anomaly | Y; N; I don't know | Radio buttons | Reported | | | x | x |
| 227. Congenital anomaly - follow-up | Follow-up / evolution of the congenital anomaly in the first 8 weeks after birth | Text | Text box | Reported | | | | x |
| **Neonatal withdrawal symptoms** | | | | | | | | |
| 228. Withdrawal symptoms | Identification of neonatal withdrawal symptoms, related to medication, drugs or alcohol use during pregnancy | Y; N; I don't know | Radio buttons | Reported | | | x | |
| 229. Withdrawal symptoms - description | Description of the neonatal withdrawal symptoms, related to medication, drugs or alcohol use during pregnancy | Text | Notes box | Reported | | | x | |
| 230. Withdrawal symptoms - duration | Duration of the neonatal withdrawal symptoms (or still ongoing) | Text | Notes box | Reported | | | x | |

| | | | | | | | | |
|---|---|---|---|---|---|---|---|---|
| 231. Withdrawal symptoms - medication | Medication used to alleviate the neonatal withdrawal symptoms | Y; N; I don't know | Radio buttons | Reported | | | x | |
| 232. Withdrawal symptoms - type of medication | Names of the medicines used to alleviate the neonatal withdrawal symptoms | Text | Notes box | Reported | | | x | |
| **Neonatal department admission** | | | | | | | | |
| 233. Admission to a neonatal department | Admission to a neonatal department or a neonatal intensive care department | My baby was not admitted to a neonatal (intensive) unit after birth; My baby was admitted to a neonatal ward after birth (N); My baby was admitted to a neonatal intensive care unit (NIC) after birth | Radio buttons | Reported | | | x | |
| 234. Neonatal department admission – reason | Reason of the admission to a neonatal department or a neonatal intensive care department | Preterm birth (< 37 weeks); Low birth weight (< 2500 grams); Oxygen deficiency at birth; Problems with breathing (specify); (congenital) problem with the metabolism (i.e., abnormal Guthrie test); Jaundice (icterus) for which phototherapy was needed; Problems with the heart (specify); An abnormally fast heart rhythm; An abnormally slow or irregular heart rhythm; Infection (specify); Problems with digestion (specify); Seizures or epileptic seizures; (severe) congenital abnormality; Stroke; Neonatal withdrawal symptoms due to the use of medication or substances during pregnancy; Problems with the temperature control; Hypotonia ; Other (specify) - Including possibility to add free text | Checkboxes with embedded text fields, Multiple choice - Notes box | Reported | | | x | |
| 235. Neonatal department - current stay | Currently staying at a neonatal department | Y/N | Radio buttons | Reported | | | x | |
| 236. Neonatal department - days | Number of days that the neonate (has) stayed at a neonatal department | Number | Text box | Reported | | | x | |
| **Feeding of the infant** | | | | | | | | |
| 237. Type of feeding | Type of feeding in the first 24 hours after birth (i.e., in the first postpartum | Breastfeeding/breast milk; Formula feeding; Mixed feeding | Radio buttons | Reported | | | x | x |

| | | | | | | | | |
|---|---|---|---|---|---|---|---|---|
| | questionnaire) or currently (i.e., in the second postpartum questionnaire) | (breastfeeding/breastmilk and formula); Other (specify) | | | | | | |
| 238. Breastfeeding/giving breast milk – since birth | Breastfeeding or giving breast milk, ever since birth | Y/N | Radio buttons | Reported | | | x | |
| 239. Breastfeeding/giving breast milk - currently | Currently breastfeeding or giving breast milk | Y/N | Radio buttons | Reported | | | x | |
| 240. Breastfeeding/giving breast milk – exclusively | Exclusively breastfeeding or giving breast milk | Y/N | Radio buttons | Reported | | | x | |
| 241. Breastfeeding/giving breast milk - duration | Duration of breastfeeding or giving breast milk | Text | Text box | Reported | | | x | |
| 242. Breastfeeding/giving breast milk - reason for cessation | The reason of breastfeeding cessation or stop giving breast milk to the infant | Text | Notes box | Reported | | | x | |
| **Neonatal complications** | | | | | | | | |
| 243. Neonatal complications | Complications or medical problems that occurred in the infant in the first 28 days after birth | Problems with breathing (specify); (congenital) problem with the metabolism (i.e., abnormal Guthrie test); Jaundice (icterus) for which phototherapy was needed; Problems with the heart (specify); An abnormally fast heart rhythm; An abnormally slow or irregular heart rhythm; Infection (specify); Problems with digestion (specify); Seizures or epileptic seizures; (severe) congenital abnormality; Cerebral haemorrhage; Neonatal withdrawal symptoms due to the use of medication or substances during pregnancy; Problems with the temperature control; Hypotonia; Other (specify) | Checkboxes with embedded text fields, Multiple choice - Notes box | Reported | | | x | x |
| 244. Neonatal hospital (re)admission | Hospital (re)admission of the infant in the first 28 days after birth (i.e., not the hospital admission for the birth itself) | Y/N | Radio buttons | Reported | | | x | x |
| 245. Neonatal hospital (re)admission - reason | Reason of the hospital (re)admission of the infant in the first 28 days after birth | Text | Notes box | Reported | | | x | x |

| | | | | | | | | |
|---|---|---|---|---|---|---|---|---|
| **246. Medication use of the neonate** | Names of the medicines used by the infant in the first 28 days after birth. If the name(s) is (are) not known, the indication can also be given. | Text | Notes box | Reported | | | x | x |

Abbreviations : EM = Pregnancy Enrolment Questionnaire; FU = Pregnancy Follow-up Questionnaire; PP 1 = First Postpartum Questionnaire; PP 2 = Second Postpartum Questionnaire.
Notes: [1]Multiplication of the neonatal outcome fields in case of plurality; all the variables of category 7 need to be competed for each live born child; [2]Structured text field with autocomplete function, linked to NATO database with country names and alpha-2 codes.

### 2 Supplementary Figure

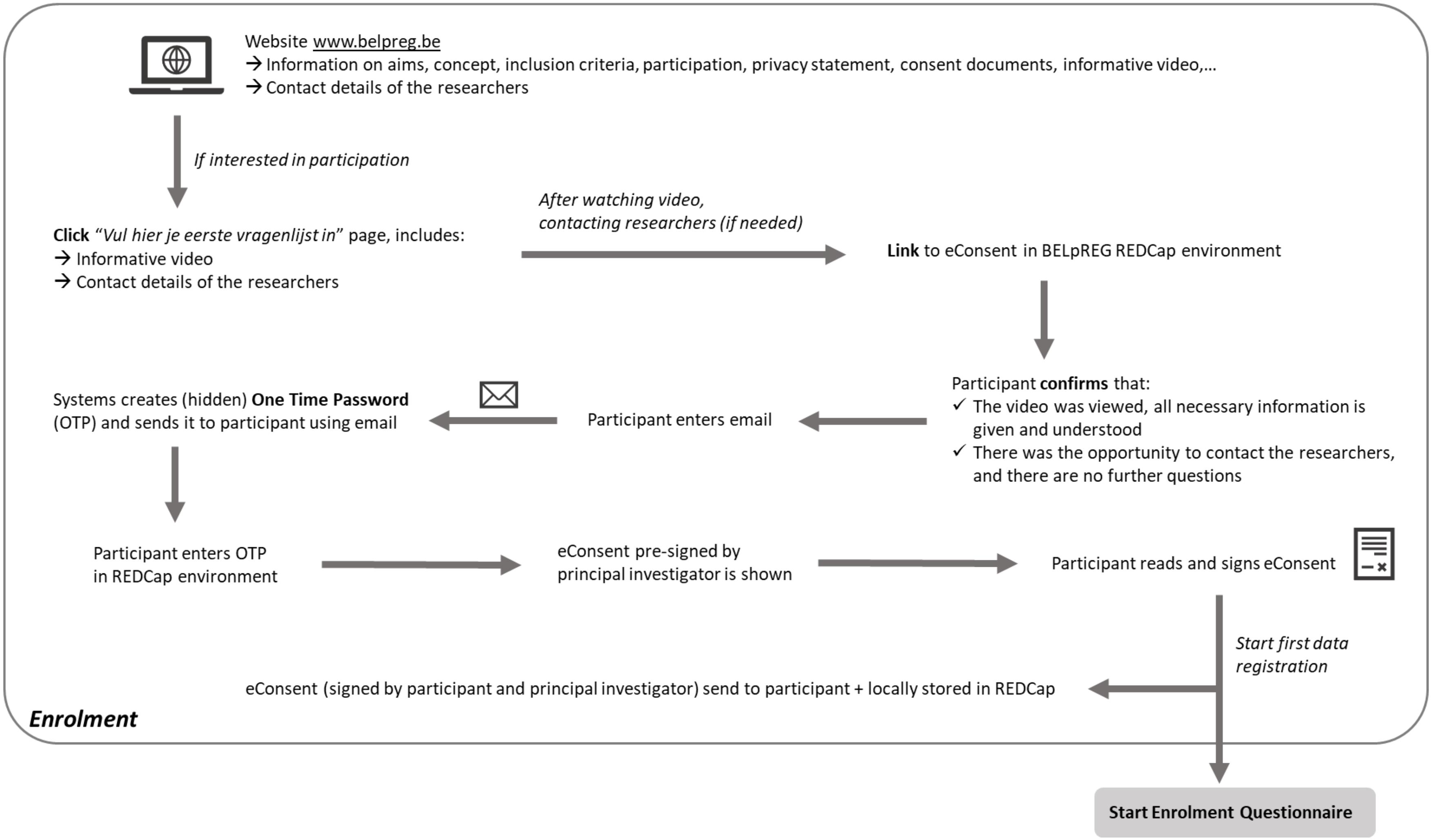

**Supplementary Figure 1.** Schematic overview of the eConsent procedure
